# Multi-season evaluation and analysis of categorical trend forecasts of influenza hospital admissions in the United States

**DOI:** 10.64898/2026.08.31.26361843

**Authors:** Jessica T. Davis, Gursharn Kaur, Annabella Hines, Michal Ben-Nun, Srinivasan Venkatramanan, Logan Brooks, Sarabeth Mathis, Marco Ajelli, Maria Litvinova, Allisandra G. Kummer, Paulo Cesar Ventura, Shreeya Mhade, David Weber, Dmitry Shemetov, Nat DeFries, Daniel J. McDonald, Teresa Yamana, Rodrigo Zepeda-Tello, Jeffrey Shaman, Rami Yaari, Sen Pei, Alexander Webber, Li Shandross, Evan Ray, Spencer Wadsworth, Jarad Niemi, William T. Redman, Luke Mullany, Richard Posner, Abhishek Mallela, Yen Ting Lin, William S. Hlavacek, Adam Smart, Amir Aman Gill, Avery Drennan, Bria Jayde Fiebiger, Ely Finn Miller, Jaechoul Lee, Joseph R. Mihaljevic, Kylie Ann Geist, Maya Baltz, Ozbej Bernik, Y-Minh B. Truong, Ye Chen, Colin James Grosvenor, Mauricio Santillana, Candice Djorno, Jiecheng Lu, Shihao Yang, Fred Lu, Leonardo Clemente, Austin G. Meyer, Clara Bay, Alessandra Urbinati, Nicolò Gozzi, Matteo Chinazzi, Minami Ueda, Nima Moghaddas, Remy LeWinter, Sara Venturini, Stefania Fiandrino, Alessandro Vespignani, Spencer J. Fox, Ehsan Suez, Mariah Salcedo, Rajath Prabhakar, B. K. M. Case, Amanda Perofsky, Cécile Viboud, James Turtle, VP Nagraj, Amy Benefield, Desiree Williams, Graham C. Gibson, Lauren Meyers, Edward Thommes, Christopher van Bommel, Rhiannon Loster, Benjamin Benteke Longaou, Monica Cojocaru, Pengfei Yue, Alexander Rodríguez, Ruipu Li, Sonika Potnis, Nicholas G. Reich, Thomas Robacker, Joseph Lemaitre, Aniruddha Adiga, Bryan Lewis, Madhav Marathe, Nibir Chandra Mandal, Stephen D. Turner, Naren Ramakrishnan, Yiqi Su, Michael Johansson, Matthew Biggerstaff, Rebecca K. Borchering

## Abstract

Forecasting influenza hospitalizations informs public health preparedness, yet questions remain about which types of forecasts best guide action. We evaluate categorical trend forecasts, which communicate probabilities of upcoming increases or decreases in epidemic trajectories, submitted to CDC’s FluSight Forecasting Challenge between Fall-2024 and Spring-2026. Teams submitted probability distributions over five categories describing direction and magnitude of week-over-week changes in laboratory-confirmed influenza hospital admissions. We assessed performance using Ranked Probability Skill Score, Brier Skill Score, and measures of forecast-observation agreement. Most models outperformed an equal-probability baseline; the FluSight ensemble ranked among the top three in the 2024–25 and 2025–26 seasons. Forecasts were most accurate during stable periods and least during periods of rapid change, with most models underestimating observed trends. Conclusions were robust to choice of scoring metric and reference model. These results support categorical trend ensembles as an approach to communicating infectious disease forecasts that may inform public health decision-making.

## 1. Introduction

In the United States, the Centers for Disease Control and Prevention (CDC) estimates that between 120,000 and 710,000 influenza hospitalizations occur each year, contributing substantially to public health burden^1^. Accurate and timely influenza forecasts can be used to mitigate this burden by providing situational awareness and informing decision-making^2–4^. Common practices for presenting influenza forecasts^5^ may be difficult to interpret and leave room for improvement in adequately communicating forecast uncertainty. Ensemble forecasts have been used as one way to simplify influenza forecasts^6,7^ by summarizing results from many forecast sources into a single source of information and often make more accurate predictions on average than forecasts made from individual models alone.^8,9^ However, there remains a need for more accurate forecasts, particularly during periods of rapid change^10,11^, as well as ways to improve communication of forecasts and their associated uncertainty broadly for public health officials and the public.

During the atypically early increase in laboratory-confirmed hospital admissions in the 2022–23 influenza season, forecasts of the magnitude of weekly influenza hospital admissions performed poorly, even once forecasts identified that increases had started ^9^. This led to a delay in public communication of influenza forecasts and motivated the development of a novel forecasting target: the probability of seeing large increases, increases, stable changes, decreases, or large decreases in the near future. CDC piloted this categorical trend target midway through the 2022–23 season and later formalized it, collecting 1- to 4-week ahead probabilistic forecasts to test whether this approach could improve forecast utility during periods of rapid change while also communicating uncertainty in the trend with probabilities that may be more directly interpretable than how forecast uncertainty is typically portrayed (e.g. prediction intervals)^12,13^. Categorical thresholds were derived from the historical distribution of week-to-week changes in influenza hospital admission rates.^14^ Thresholds were first based on FluSurv-NET^15,16^, since the National Healthcare Safety Network(NHSN)^17^ did not yet cover a complete influenza season; as more NHSN laboratory confirmed influenza hospital admissions data became available, thresholds were updated using both combined datasets.

Evaluating probabilistic forecasts of categorical observations is a well-studied task that spans diverse fields including geopolitics, climate science, economics, machine learning and more recently in forecasts of respiratory pathogens in Europe^18^. In geopolitical forecasting, large-scale initiatives such as the Good Judgment Project^19^ and Intelligence Advanced Research Projects Activity Aggregative Contingent Estimation tournaments^20^ have assessed the prediction accuracy of binary and multi-choice targets. Additional approaches have been developed within meteorology, climate sciences, and space weather for evaluating forecasts of ordered categories like ranges of temperature and precipitation or solar flare predictions.^21–23^ For macroeconomic indicators and energy load forecasting, similar approaches are used to evaluate full probability distributions of outcomes such as inflation, Gross Domestic Product growth, and discretized electricity price ranges.^24,25^

Drawing on this breadth of approaches, we leverage established scoring methods and forecast data from the largest, longest-standing infectious disease forecast collaboration to evaluate novel categorical forecasts of per capita influenza hospital admission trends. We describe these efforts in detail and explore how forecast performance differs systematically through time, location, by horizon, and observed trend category, including whether forecasts show systematic errors toward over-or under-estimating the observed trend. We also compare the performance of these categorical forecasts to analogous probabilities derived from FluSight ensemble forecasts of hospital admission counts and assess the robustness of these findings to inform whether and how categorical forecasts should be collected and communicated publicly. Here, we focus primarily on the 2024–25 and 2025–26 influenza seasons, as these probabilistic forecasts were generated using the same definitions for each category.

## 2. Results

We evaluate categorical trend forecasts of changes in laboratory-confirmed influenza hospital admission rates submitted to CDC’s FluSight Challenge during the 2024–25 and 2025–26 influenza seasons. Forecasts were submitted for 52 jurisdictions (50 U.S. states, Washington D.C., Puerto Rico), and nationally, corresponding to observed 1– to 4–week ahead changes in per capita influenza hospital admissions. Each forecast assigned probabilities to five trend categories: *large decrease*, *decrease*, *stable*, *increase*, and *large increase*, defined by thresholds on the rate of change in weekly laboratory-confirmed influenza hospital admissions (see Methods). Each season consisted of 27 submission weeks, spanning from November 23, 2024, to May 31, 2025, (excluding January 25, 2025, due to a delayed data release) and from November 22, 2025, to May 23, 2026. Details on the underlying surveillance data and model inclusion criteria are provided in the Supplemental Information (SI).

### 2.1 Observed categorical trends

The 2024–25 and 2025–26 influenza seasons differed substantially in their epidemic characteristics. These differences are visible in one-to four-week ahead categorical trends in laboratory-confirmed influenza hospital admission rates (Figs. 1, S1).

**Figure 1:**
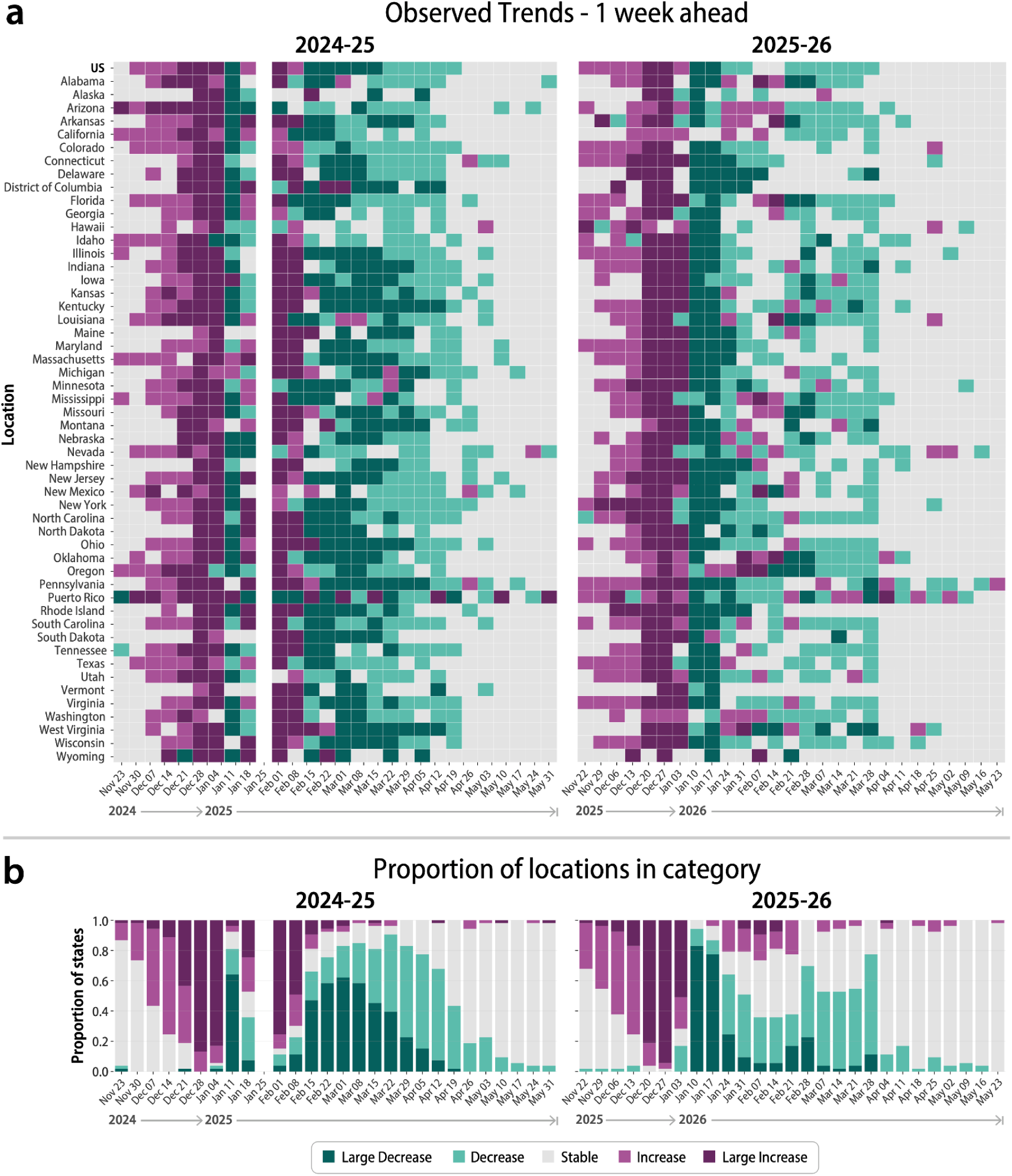
Observed categorical one-week ahead trends in the 2024–25 and 2025–26 seasons. (a) One-week ahead changes observed for each jurisdiction and the nation each week within the 2024–25 and 2025–26 influenza seasons. Data from the week of January 25, 2025 are missing due to delays in reporting that prevented models from submitting regularly scheduled forecasts. This week was dropped from the analysis, even though observational data was eventually released. (b) The proportion of locations (all jurisdictions and nationally) which were assigned to each category during a given week in the 2024–25 and 2025–26 influenza seasons.

The 2024–25 season was classified by the CDC as high severity, the first since the 2017–18 season.^26^ Nationally, laboratory-confirmed influenza hospital admissions surpassed 1000 per week by November 2024 and rose to over 39,000 in the first week of January 2025. The season was dominated by influenza A(H1N1) and influenza A(H3N2) viruses, which co-circulated at roughly equal levels.^27^ There were two periods of increasing activity, beginning the weeks of December 7, 2024, and February 1, 2025, where *large increases* and *increases* were observed in more than 50% of jurisdictions. These two increasing periods were separated by at least one week in which more than 80% of jurisdictions observed a *decrease* or *large decrease* (Fig. 1a). Starting the week of February 15, 2025, *decrease or large decrease* were the most frequently observed categories, until April 19, 2025, after which *stable* became the most commonly observed category through the end of the evaluation period (Fig. 1b).

In contrast, the 2025–26 influenza season was classified as moderate severity overall^28^. Influenza hospital admissions rose sharply and synchronously across the United States during late November to December 2025. During the weeks of December 20 and December 27, 2025, all but one jurisdiction observed an *increase* or *large increase* in one-week ahead trends. By the week of January 10, 2026, 51 of 53 jurisdictions were classified as a *decrease* or *large decrease*. A rise in influenza B virus circulation beginning in February 2026, slowed the decline in some jurisdictions, shifting them from *decrease* to *stable* classifications (Fig. 1b). Stable trends were not observed consistently across the U.S. until April 2026.

Overall, the distribution of observed categories was similar across seasons, with stable being the most commonly observed category in both the 2024-25 (32.1%) and 2025-26 (32.6%) seasons. Differences arose in the balance of increases versus decreases and in the frequency of large decrease and large increase categories, reflecting the underlying seasonal dynamics (Table S2). Category frequencies depend on the threshold definitions used to classify trends. As an illustration, Section S1.2 compares the observed categories for the 2023–24 season under both the original 2023–24 thresholds, which were and the 2024–25/2025–26 thresholds, showing that the narrower stable-category thresholds used in later seasons led to fewer observations being classified as stable (Fig. S2).

### 2.2 Quantifying forecast performance

We evaluated categorical trend forecasts from 20 models in 2024–25 and 16 in 2025–26. We excluded additional models that did not meet the inclusion criteria (Sec. S2). In 2024–25, 15 of the 20 and in 2025–26, 11 of the 16 evaluated models were contributed by participating teams, with the remainder of models consisting of FluSight ensembles (built from contributed forecast submissions) and reference models (see Methods Sec 4.3). Similar to the count-based forecasts, model methodologies spanned a wide range of approaches, including, mechanistic compartmental models, statistical time series approaches, and machine learning models.^13^ We solicited 5,724 categorical forecast targets (unique submission date, jurisdiction, and week ahead combinations) in both the 2024–25 and 2025–26 seasons (submission frequency varied by team, Table S5).

Each week during the forecast season, the CDC FluSight team constructed the categorical ensemble, referred to as the FluSight-ensemble, in which the forecast probabilities for each category were averaged across those of all eligible submitted model forecasts. For this analysis, we also calculated categorical forecast probabilities from the probability distributions associated with the count-based FluSight ensemble forecasts (FluSight-ens_q_cat). We additionally constructed FluSight-ens_q_cat_sub using the same approach but restricted inclusion to the subset of count-based models that also contributed to the categorical FluSight-ensemble (see Table 3).

#### 2.2.1 Agreement between forecasts and observed categories

As an initial assessment of forecast quality, we compared how often a model’s highest-probability category matched the observed trend in the 1-week ahead predictions for all jurisdictions and nationally. Each forecast was classified as a *primary match* if the highest-probability category matched the observed category, including ties for the highest probability, a *secondary match* if the second-highest distinct probability corresponded to the observed category, and *not a match* otherwise. Secondary matches were included to provide additional context for forecasts without a primary match, distinguishing cases in which the observed category was the next most probable outcome from those with greater disagreement. Across submitted models, primary match percentages for 1-week-ahead forecasts ranged from 34.9% to 60.4% in 2024–25 and from 50.8% to 66.7% in 2025–26 (Table S6). Correspondingly, the proportion of forecasts classified as no match ranged from 17.1% to 51.6% in 2024–25 and from 14.3% to 32.6% among submitted models in 2025–26. The FluSight ensemble ranked among the top-performing models in both seasons, achieving the highest 1 week-ahead primary-match percentage (60.4%) in 2024–25 and the third highest (63.3%) in 2025–26.

In 2024–25, the majority (75%) of 1-week ahead forecasts from all submitted models that were not a match occurred between December 21, 2024, and February 15, 2025, coinciding with periods of increasing laboratory-confirmed influenza hospital admissions (Fig. 2). In 2025–26, the weeks of December 20, 2025, and January 10, 2026, accounted for 13% and 14% of all seasonal 1-week ahead forecast mismatches, respectively. During those weeks, 56% and 60% of forecasts were classified as no match, compared with a maximum of 37% during any other week. These weeks also coincided with the highest proportions of observed *large increase* and *large decrease* categories during the seasons (Fig. 1).

**Figure 2:**
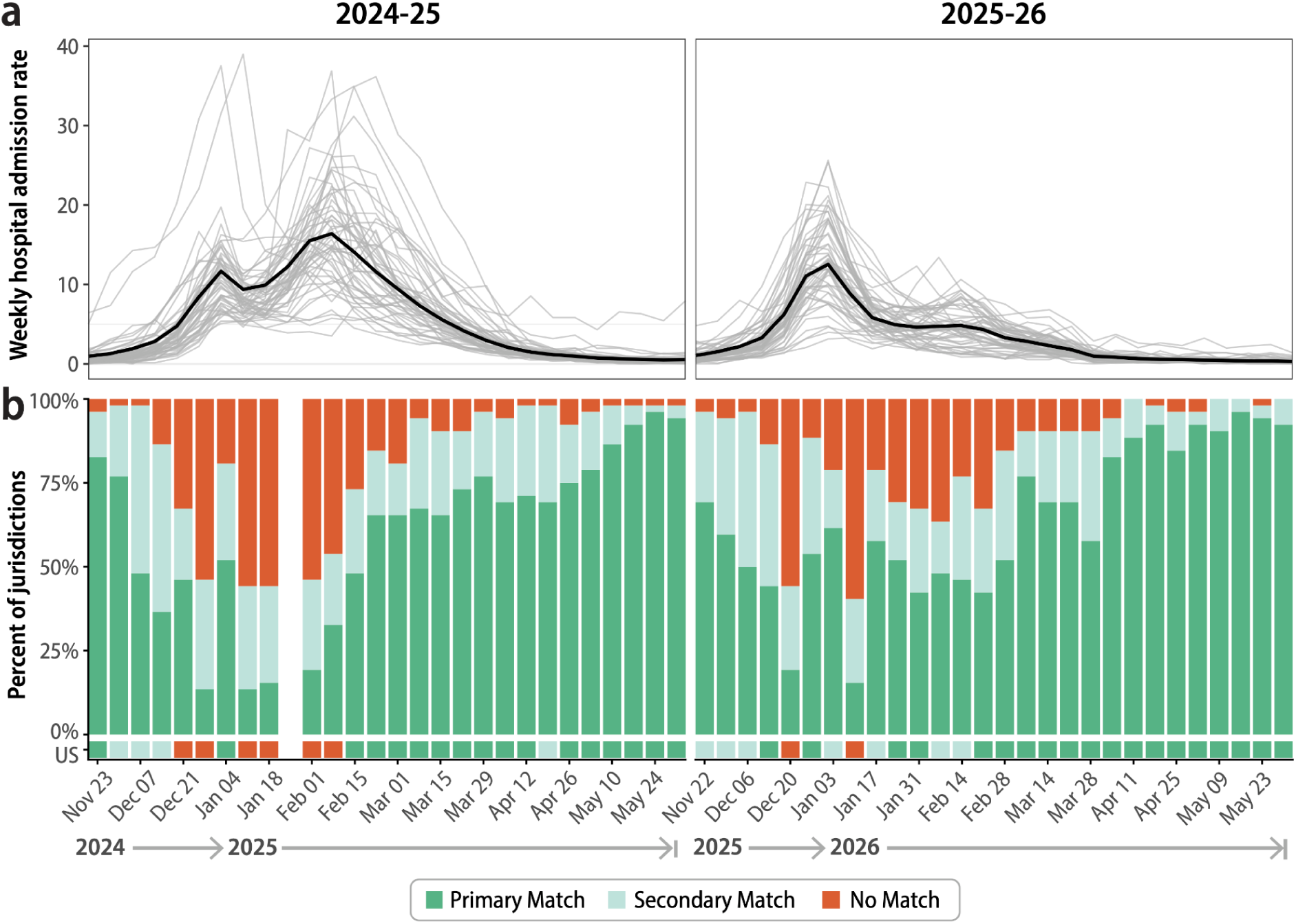
Observed weekly laboratory-confirmed influenza hospital admissions and corresponding matches or mismatches with forecasted one-week categorical trends. (a) Epidemic curves for the weekly observed laboratory-confirmed influenza hospital admission rates in the 2024–25 (left) and 2025–26 (right) seasons. Each gray line represents a single jurisdiction. The national rate is shown as a black line. (b) The proportion of jurisdictions within each week that the categorical ensemble (FluSight-ensemble) correctly or incorrectly predicted the observed one-week trend category. Primary Match indicates that the categorical ensemble assigned the highest probability to the observed category. Secondary Match indicates that the categorical ensemble assigned the second highest probability to the observed category. Predictions are considered No Match if neither the first nor second highest probabilities are assigned to the observed category.

Focusing on the FluSight-Ensemble, the frequency of agreement between the observed trend and predictions varied across both seasons (1-week ahead changes in Fig. 2, and 2-4-week ahead changes in Fig. S3). Across all horizons and jurisdictions, the ensemble achieved a primary match for 59.8% of forecasts in 2024–25 and 62.7% in 2025–26. At the national level, the categorical ensemble correctly predicted the 1-week ahead rate-trend category by assigning the highest probability to the observed category 65% of the time (35 weeks), the second highest probability 20% (11 weeks), and attributed the wrong directional trend 15% (8 weeks) across both seasons. The proportion of matched forecasts decreased during periods of increasing influenza hospital admissions in each season, particularly for 3- and 4-week ahead changes (Fig. S3). Following the seasonal peak, ensemble forecast performance improved across all horizons as influenza activity stabilized and declined, with the majority of jurisdiction forecasts classified as either a primary or secondary match.

#### 2.2.2 Model and ensemble skill within and across seasons

As part of a more formal evaluation framework, we also assess forecast performance using the Brier score^29^, a common probabilistic score for categorical forecasts. We convert this into a skill score (the Brier skill score, BSS) to measure the improvement in performance relative to a reference model, where positive values indicate better performance than the reference model and negative values indicate worse performance (Table 1). Our primary reference model is FluSight-equal_cat, which assigns equal probability to each of the five trend categories (see Sec. 4.4 for details).

**Table 1:**
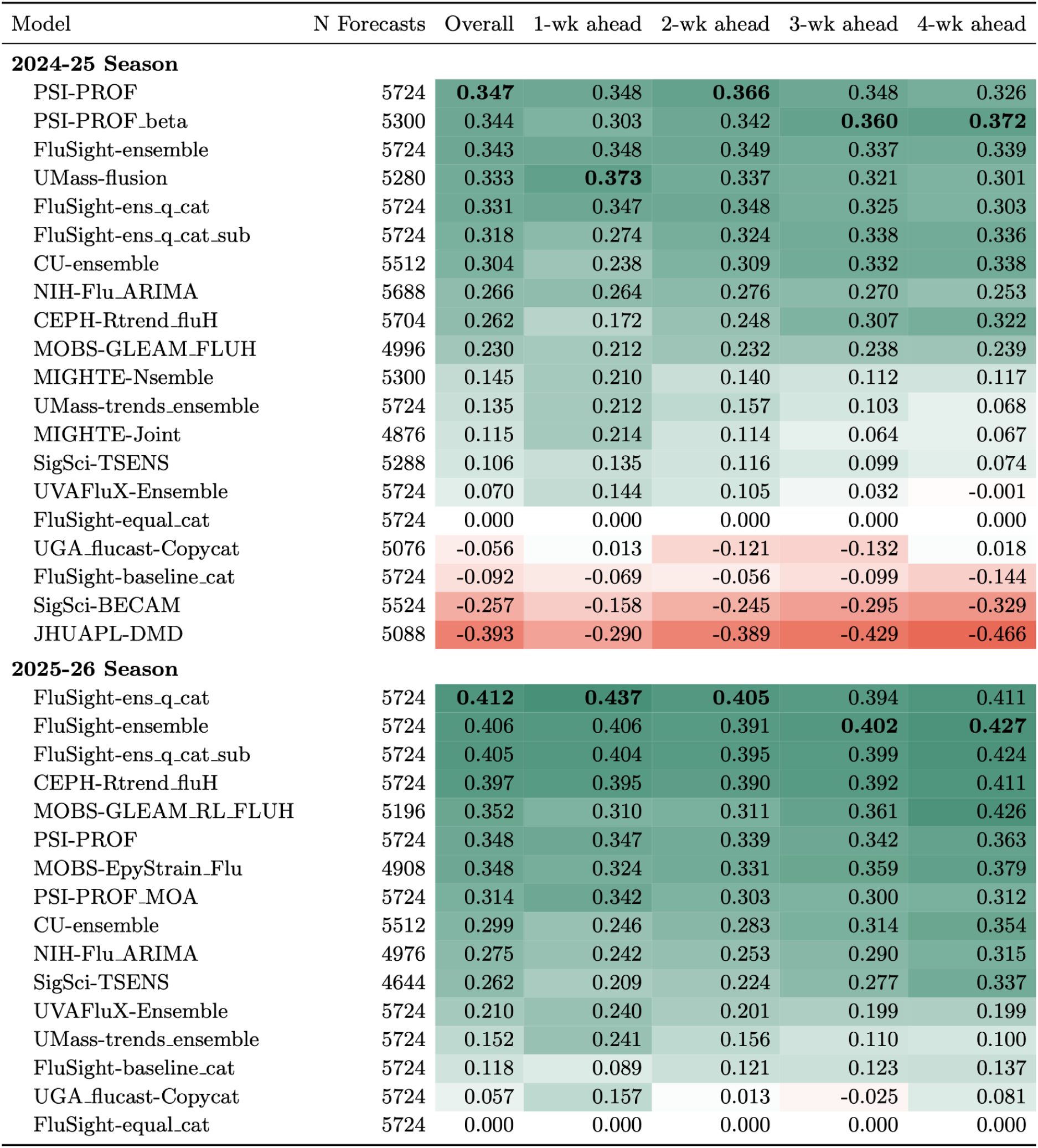
Brier skill scores (BSS) for each model aggregated across all individual forecast targets by season (overall) and separately for 1- to 4- week ahead change forecasts within each season. Skill scores are calculated with FluSight-equal_cat as the reference model. A BSS> 0 indicates that the model performed better than the reference model and BSS< 0 denotes worse performance. The highest skill score in each column for each season is shown in bold.

| Model | N Forecasts | Overall | 1-wk ahead | 2-wk ahead | 3-wk ahead | 4-wk ahead |
| --- | --- | --- | --- | --- | --- | --- |
| **2024-25 Season** |  |  |  |  |  |  |
| PSI-PROF | 5724 | **0.347** | 0.348 | **0.366** | 0.348 | 0.326 |
| PSI-PROF_beta | 5300 | 0.344 | 0.303 | 0.342 | **0.360** | **0.372** |
| FluSight-ensemble | 5724 | 0.343 | 0.348 | 0.349 | 0.337 | 0.339 |
| UMass-flusion | 5280 | 0.333 | **0.373** | 0.337 | 0.321 | 0.301 |
| FluSight-ens_q_cat | 5724 | 0.331 | 0.347 | 0.348 | 0.325 | 0.303 |
| FluSight-ens_q_cat_sub | 5724 | 0.318 | 0.274 | 0.324 | 0.338 | 0.336 |
| CU-ensemble | 5512 | 0.304 | 0.238 | 0.309 | 0.332 | 0.338 |
| NIH-Flu_ARIMA | 5688 | 0.266 | 0.264 | 0.276 | 0.270 | 0.253 |
| CEPH-Rtrend_fluH | 5704 | 0.262 | 0.172 | 0.248 | 0.307 | 0.322 |
| MOBS-GLEAM_FLUH | 4996 | 0.230 | 0.212 | 0.232 | 0.238 | 0.239 |
| MIGHTE-Nsemble | 5300 | 0.145 | 0.210 | 0.140 | 0.112 | 0.117 |
| UMass-trends_ensemble | 5724 | 0.135 | 0.212 | 0.157 | 0.103 | 0.068 |
| MIGHTE-Joint | 4876 | 0.115 | 0.214 | 0.114 | 0.064 | 0.067 |
| SigSci-TSENS | 5288 | 0.106 | 0.135 | 0.116 | 0.099 | 0.074 |
| UVAFluX-Ensemble | 5724 | 0.070 | 0.144 | 0.105 | 0.032 | -0.001 |
| FluSight-equal_cat | 5724 | 0.000 | 0.000 | 0.000 | 0.000 | 0.000 |
| UGA_flucast-Copycat | 5076 | -0.056 | 0.013 | -0.121 | -0.132 | 0.018 |
| FluSight-baseline_cat | 5724 | -0.092 | -0.069 | -0.056 | -0.099 | -0.144 |
| SigSci-BECAM | 5524 | -0.257 | -0.158 | -0.245 | -0.295 | -0.329 |
| JHUAPL-DMD | 5088 | -0.393 | -0.290 | -0.389 | -0.429 | -0.466 |
| **2025-26 Season** |  |  |  |  |  |  |
| FluSight-ens_q_cat | 5724 | **0.412** | **0.437** | **0.405** | 0.394 | 0.411 |
| FluSight-ensemble | 5724 | 0.406 | 0.406 | 0.391 | **0.402** | **0.427** |
| FluSight-ens_q_cat_sub | 5724 | 0.405 | 0.404 | 0.395 | 0.399 | 0.424 |
| CEPH-Rtrend_fluH | 5724 | 0.397 | 0.395 | 0.390 | 0.392 | 0.411 |
| MOBS-GLEAM_RL_FLUH | 5196 | 0.352 | 0.310 | 0.311 | 0.361 | 0.426 |
| PSI-PROF | 5724 | 0.348 | 0.347 | 0.339 | 0.342 | 0.363 |
| MOBS-EpyStrain_Flu | 4908 | 0.348 | 0.324 | 0.331 | 0.359 | 0.379 |
| PSI-PROF_MOA | 5724 | 0.314 | 0.342 | 0.303 | 0.300 | 0.312 |
| CU-ensemble | 5512 | 0.299 | 0.246 | 0.283 | 0.314 | 0.354 |
| NIH-Flu_ARIMA | 4976 | 0.275 | 0.242 | 0.253 | 0.290 | 0.315 |
| SigSci-TSENS | 4644 | 0.262 | 0.209 | 0.224 | 0.277 | 0.337 |
| UVAFluX-Ensemble | 5724 | 0.210 | 0.240 | 0.201 | 0.199 | 0.199 |
| UMass-trends_ensemble | 5724 | 0.152 | 0.241 | 0.156 | 0.110 | 0.100 |
| FluSight-baseline_cat | 5724 | 0.118 | 0.089 | 0.121 | 0.123 | 0.137 |
| UGA_flucast-Copycat | 5724 | 0.057 | 0.157 | 0.013 | -0.025 | 0.081 |
| FluSight-equal_cat | 5724 | 0.000 | 0.000 | 0.000 | 0.000 | 0.000 |

Overall, the average Brier skill scores were higher in 2025–26 than in 2024–25: 12 of 15 submitted models outperformed the equal-categorical reference model in 2024–25, and all models outperformed it in 2025–26. Skill scores varied within each season (Fig. 3), generally declining during periods of increasing influenza hospital admissions at the beginning of each season and near seasonal peaks (Fig. 3). Many models received negative BSS values for substantial portions of each season, indicating performance worse than the uninformative, equal probability model. In the 2024–25 season, nearly all model forecasts received negative BSS values during late December 2024 and late January 2025, coinciding with the two periods of increasing influenza hospital admissions (Figs. 2a, c and 4a). In the 2025–26 season, we observe a similar pattern, with BSS declines starting in mid-December 2025, though to a lesser extent (Fig. 3b).

**Figure 3:**
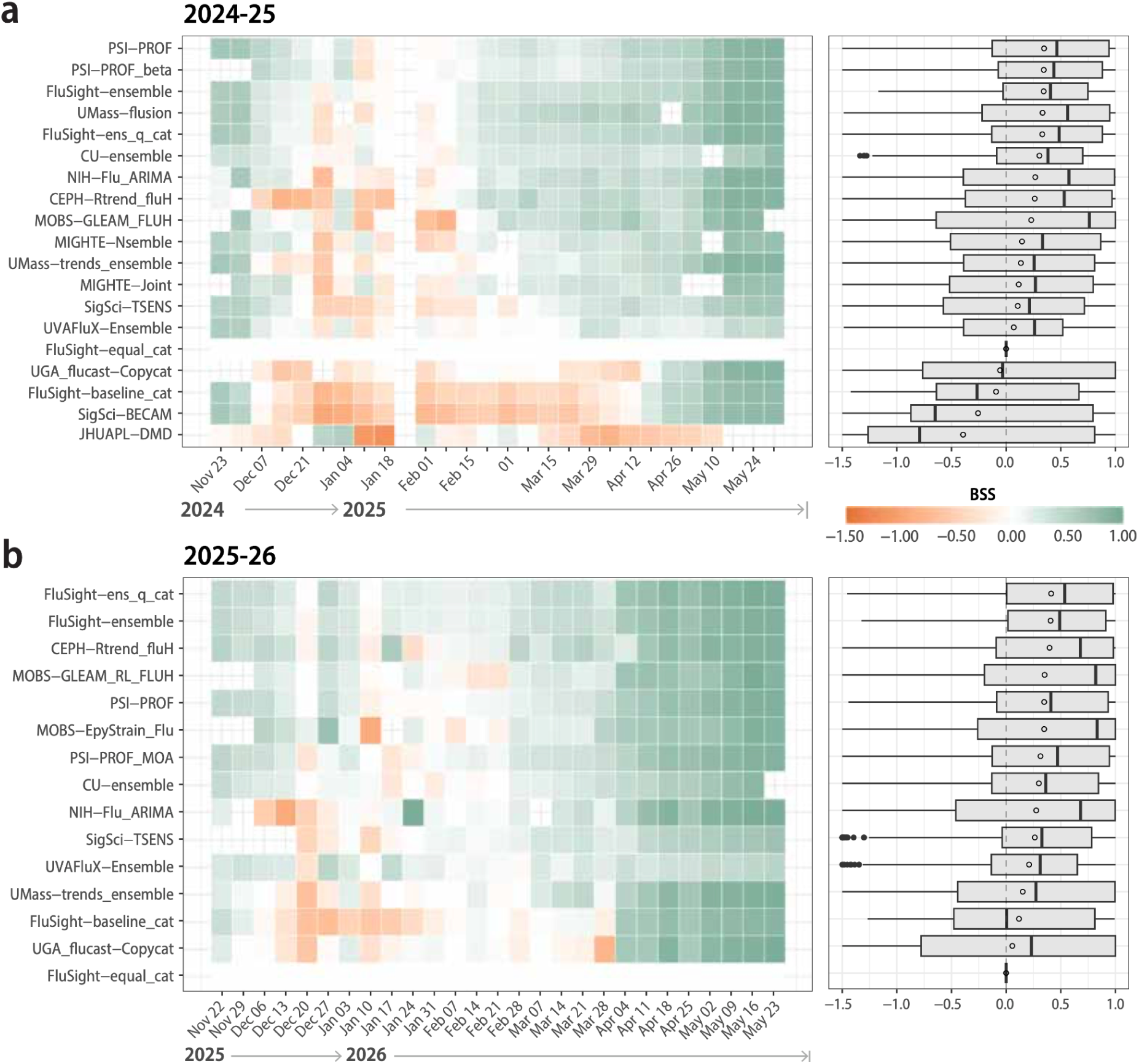
Model specific BSS across evaluated influenza seasons. Weekly mean BSS for evaluated models averaged over jurisdictions where both the model and the reference model (FluSight-equal_cat) are available (left) and the aggregated distribution of the BSS across jurisdictions and forecast weeks (right) for the 2024-25 (a) and 2025–26 (b) seasons. The boxes represent the interquartile range (IQR) of the BSS distribution, the whiskers (lines extending the box) are 1.5x the IQR and points beyond whiskers denote outliers. Circles indicate the overall BSS, in this case the mean, and the solid vertical lines represent the median BSS.

**Figure 4:**
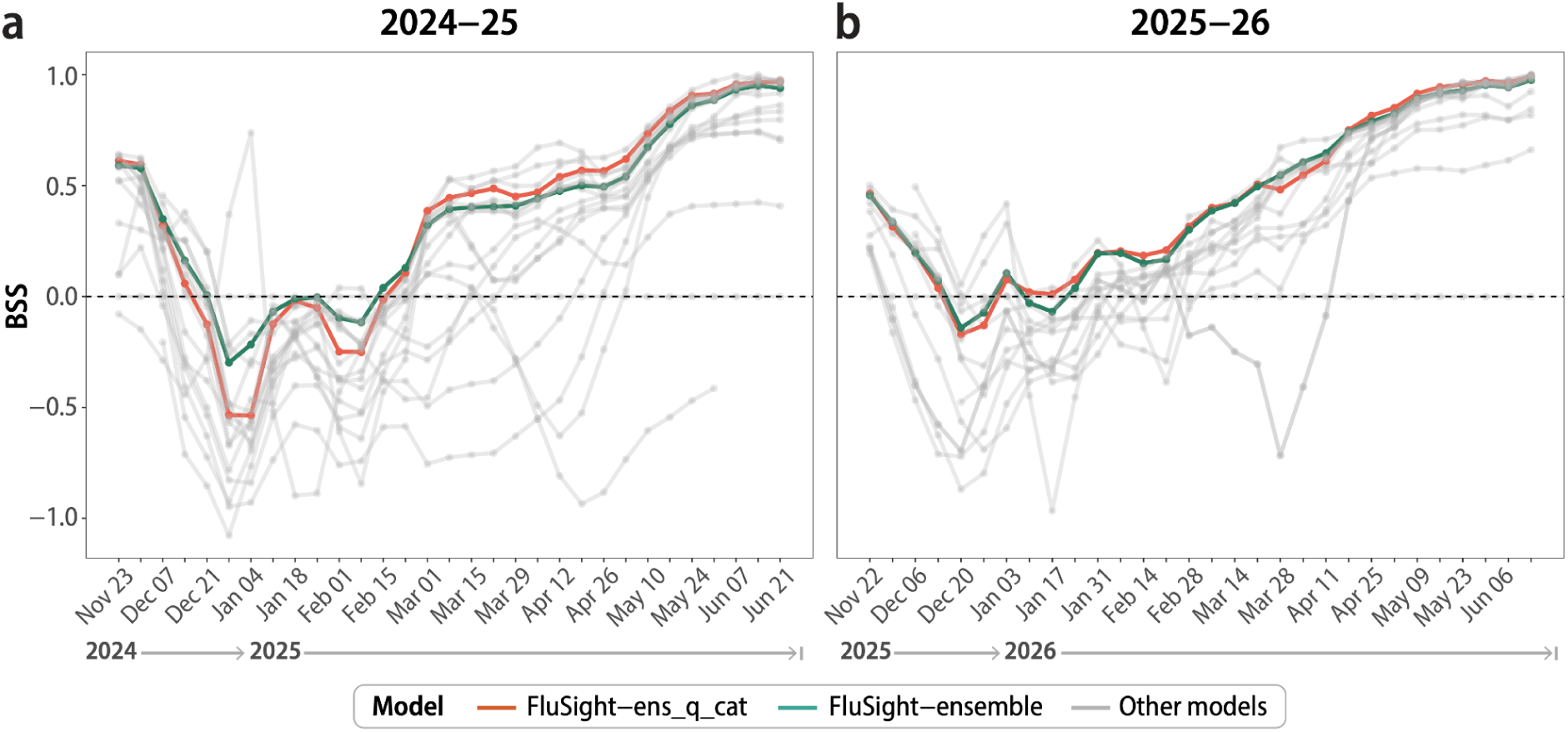
Temporal model specific performance during evaluated influenza seasons. Performance of categorical forecasts of trends in influenza hospital admissions in 2024-25 (a) and 2025-26 (b). Forecast model performance is indicated by the average BSS for each model across all locations and across all 1- to 4-week ahead change forecasts for each forecast week. BSS values for contributed models are indicated by gray lines. BSS scores for FluSight-ens_q_cat and FluSight-ensemble are shown in orange and green respectively. BSS values are computed in reference to FluSight-equal_cat. Values less than zero indicate that the equal probability reference model outperfomed the corresponding model, while values greater than zero indicate better performance.

The FluSight ensemble ranked 3rd out of 20 models in 2024–25 and 2nd out of 16 in 2025–26 by aggregated BSS (Table 1). However, performance varied across jurisdictions (Fig. S4). In 2024–25, the mean FluSight-ensemble BSS across jurisdictions ranged from 0.226 to 0.491 (median 0.347; IQR 0.320–0.387), with the lowest values in Hawaii, Louisiana, and Rhode Island and the highest values in South Dakota (Fig. S4 and Fig. S5). In the 2025–26 season, the mean BSS ranged from 0.159 to 0.596 (median 0.404; IQR 0.367–0.465), with the lowest values in Puerto Rico, Nevada and Pennsylvania, and the highest in the District of Columbia (Fig. S4).

As an alternative approach to generating categorical trend probabilities, we compared the FluSight-ensemble with another ensemble, FluSight-ens_q_cat, built by converting the count-based ensemble of weekly laboratory-confirmed influenza hospital admissions, the original FluSight forecasting target, into categorical trend probabilities (Sec. 4.3, Table 3). Agreement between the two approaches was high, with the models assigning the highest probability to the same category in 84% of forecasts in 2024–25 and 87% in 2025–26. Relative performance varied by season (Fig. 4). In 2024–25, the FluSight-ensemble outperformed FluSight-ens_q_cat through mid-February, roughly the growth-and-peak period for most locations, before the rank switched later in the season (Fig. 4a). In 2025–26, the two ensembles performed similarly throughout the season (Fig. 4b).

We found that the slight differences in performance between the two ensembles were not attributable to differences in the number of models incorporated. FluSight-ens_q_cat aggregates forecasts from more models than the FluSight-ensemble (46 in 2024–25, 55 in 2025–26) since not all models that contribute count-based forecasts to FluSight also contribute categorical forecasts. Restricting the alternative ensemble to aggregate forecasts only from the models contributing to the FluSight-ensemble (FluSight-ens_q_cat_sub) left the two ensembles within two rank positions of each other in both seasons. Few individual models outperformed either ensemble in both seasons, and none did so consistently (Table 1).

Given the similarity in categorical and count-based performance for the two ensembles, we tested whether this relationship held more generally. We calculated the per capita weighted interval score (WIS) on count-based forecasts to control for population size and expressed these relative to the FluSight-baseline model (relative WIS). We then examined the rank correlation between relative WIS and the corresponding BSS of each model. We found a moderate to strong correlation between these values in both seasons (Spearman’s r = 0.81 in 2024–25 and r = 0.58 in 2025–26; Fig. S6), indicating that models performing well for the categorical trend target also tended to perform well for the original count-based target.

As an additional check on model performance, we quantified the impact of changes and updates to the surveillance data. Categorical forecasts are defined relative to recent observations, and since those observations can change with data updates, the observed category between initial reporting and final revision can differ. We examined whether the magnitude of these data revisions was associated with model performance and found no consistent association in either season (SI Sec. S6).

#### 2.2.3 Sensitivity of evaluation to the reference model and scoring metric

In the previous section, model performance was assessed using the BSS, which makes specific assumptions about which reference model measures “skill” and how errors are penalized across categories. To test the sensitivity of our results to these assumptions, we further examine the impact on model performance of two choices: the reference model and the scoring rule. We consider an alternative to the FluSight-equal_cat reference model: the FluSight-baseline_cat model, in which categorical probabilities are derived from a flat baseline forecast with uncertainty estimated from historical week-to-week changes^30^ (see Sec. 4.3). This is the standard baseline model generated in the FluSight Challenge for count-based forecasts. We observe that models generally have higher skill scores when evaluated against the FluSight-baseline_cat compared to FluSight-equal_cat (Figs. 5a, S11). This reflects the fact that most models better capture directional change than the FluSight-baseline_cat, whose trend probabilities are nearly symmetric between increasing and decreasing trends. However, during time periods when the trends are classified as stable, the FluSight-baseline_cat model is more difficult to outperform because it concentrates probability mass near *stable*. Despite these differences, overall skill scores under the two reference models were highly correlated in both the 2024–25 and 2025–26 seasons (Fig. 5a, S11).

**Figure 5:**
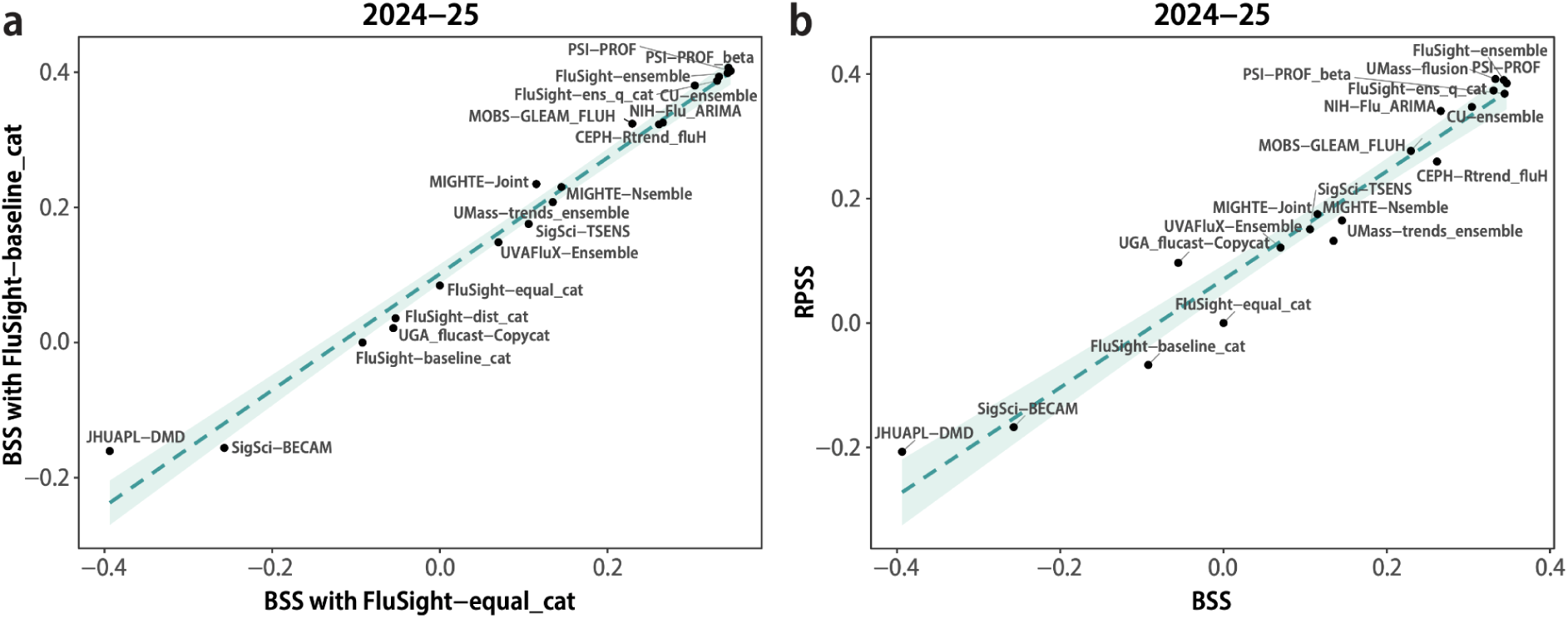
Sensitivity analysis of reference model and scoring metric. (a) Comparison of BSS across reference models. (b) Comparison of BSS and RPSS using the equal-probability reference model (FluSight-equal_cat) for all eligible models during 2024–25. The dashed line indicates the linear regression fit and the blue shaded region represents the associated 95% confidence interval.

To evaluate sensitivity to the choice of scoring rule, we compared the BSS against the skill score of the ranked probability score (RPS). We use the Brier score as our primary metric throughout the main text because it is straightforward to interpret and communicate. However, it penalizes any incorrect category equally regardless of how far it is from the observed outcome. Alternatively, the RPS accounts for the ordinal structure of the trend categories by comparing the cumulative distribution of forecast probabilities with that of the observed outcome across ordered categories, penalizing forecasts more when they are farther from the observed category (see Methods 4.4). As with BSS, skill under RPS, the ranked probability skill score (RPSS), is measured relative to a reference model. Here we compare RPSS to BSS using FluSight-equal_cat as the reference for both. Model performance and relative rankings were largely consistent between the two metrics (Fig. 5b for 2024–25, Fig. S12 for 2025–26).

### 2.3 Directional bias in categorical forecasts

While skill scores summarize performance in a single number, they are unable to reveal whether errors follow a systematic pattern. For example, do models tend to overestimate (e.g., forecast an increase when the observed trend was stable or a decrease) or underestimate (e.g., forecast a decrease when the observed trend was stable or an increase) predictions. We evaluate these potential systematic errors by constructing probability-weighted, multi-category confusion matrices for each team across both seasons. Each element (*i*, *j*) in a matrix represents the normalized, forecasted probability mass assigned to category *i* when the true observed category was *j*, aggregated across all forecasted weeks ahead, submissions, and locations, and renormalized such that each column sums to one. In Fig. 6a, we highlight the FluSight-ensemble, aggregating the five trend categories into *increase* (summing the probabilities of the *increase* and *large increase* categories), *stable*, and *decrease* (summing the probabilities of the *decrease* and *large decrease* categories) trends for readability (all models reported in Fig. S7 and Fig. S8). The diagonal of the matrix (top left to bottom right) represents the probability mass assigned to the correct category when the forecasted and observed categories matched. Cells above and below the diagonal represent probability mass assigned to incorrect categories, reflecting underestimation or overestimation of the observed trend, respectively. Models were most accurate at identifying stable periods, with 17 of 18 assigning the highest probability to the correct category when the observed trend was stable in 2024–25 and all 14 of 14 in 2025–26 (excluding the two reference models).

**Figure 6:**
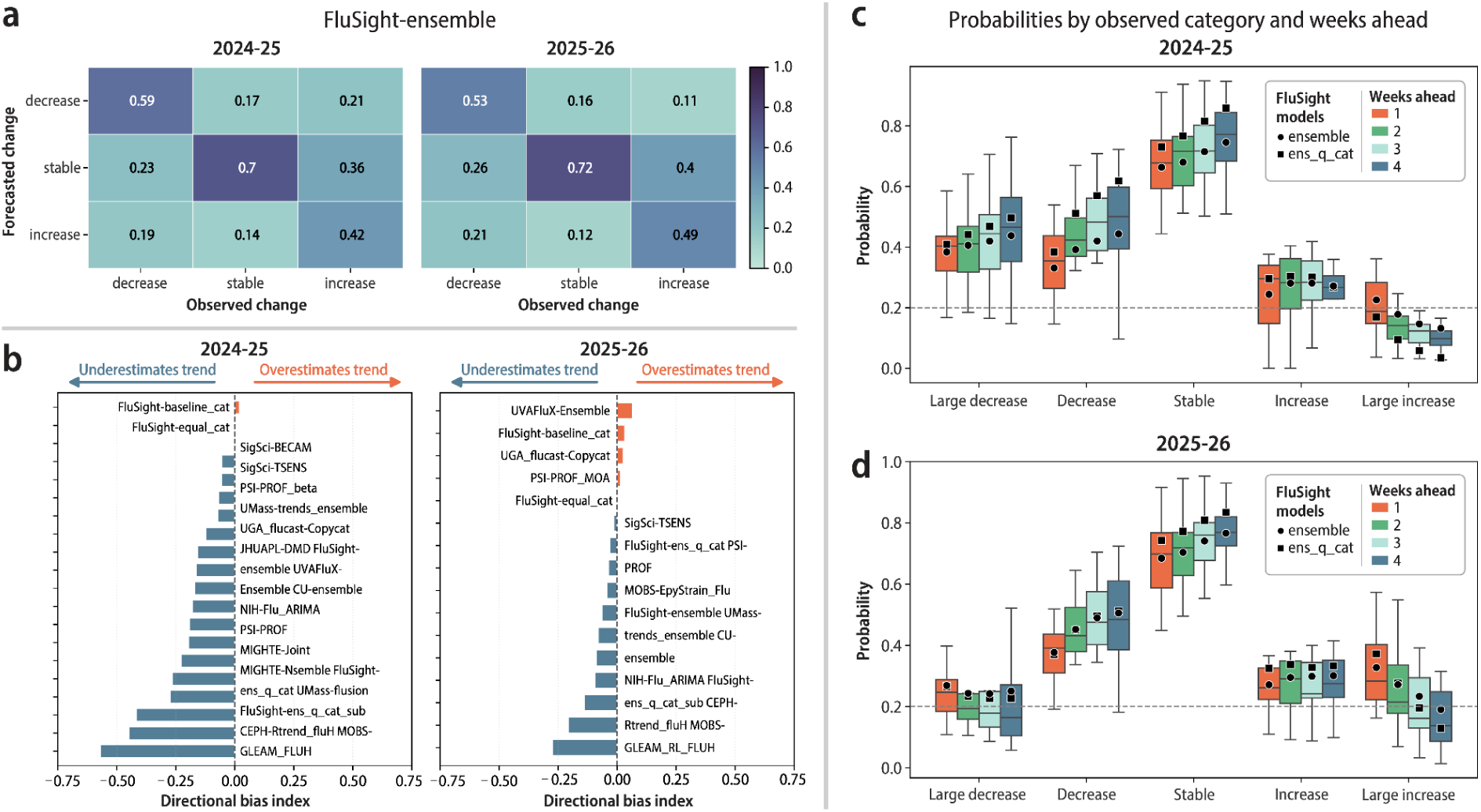
Categorical trend forecast classification, directional bias, and calibration across the 2024–25 and 2025–26 influenza seasons. (a) Confusion matrices for the categorical FluSight ensemble, showing the probability mass associated to each observed/forecasted category pair for the 2024–25 (left) and 2025–26 (right) seasons. Forecast categories were collapsed into three classes (decrease, stable, increase) by summing predicted probabilities across the two decrease bins and the two increase bins, respectively; (b) the directional bias for each included model in the 2024–25 (left) and 2025–26 (right) seasons, computed from each model’s full confusion matrix. Identical model names across seasons do not imply that the exact same forecasting methodology was used. (c–d) Distribution of the accuracy metric (the probability assigned to the observed category) across forecasting teams, split by observed categorical outcome (large decrease, decrease, stable, increase, large increase) and forecast horizon (1–4 weeks ahead), for the 2024–25 (c) and 2025–26 (d) seasons, aggregated across submission weeks and jurisdictions. The two FluSight categorical models (FluSight-ensemble and FluSight-ens_q_cat) are overlaid as distinct markers; the dashed horizontal line at 0.2 marks the accuracy value expected by chance under a uniform five equal category assignment.

Accuracy was lower and asymmetric for directional trends, as models identified decreases far more reliably than increases. We can better quantify this directional error by looking at where the probability mass falls when model predictions are incorrect (i.e. above or below the diagonal). We refer to this as the directional bias of the model, calculated as the difference between the probability mass below the diagonal, where modeling teams overestimate the trend (e.g., forecast an increase when the observed trend was a decrease), and above the diagonal, where modeling teams underestimate the trend (e.g., forecast a decrease when the observed trend was an increase), divided by the total probability mass not on the diagonal (Fig. 6b, Sec. S3.4). This quantity ranges from-1 (all off-diagonal mass underestimates the trend) to +1 (all overestimates), with 0 indicating symmetric errors. This quantity was negative across nearly all models in both seasons, indicating a systematic tendency to underestimate trends.

In addition to demonstrating directional bias, the confusion matrices can also be used to summarize overall model accuracy. Using the diagonal of the confusion matrices, we can extract a probability-weighted accuracy measure for each team. However, although this metric moderately correlates with the BSS, across teams, seasons, and forecasted weeks ahead (Fig. S9), we will treat this measure as a descriptive type of error, rather than a way to rank model performance as it is not a proper score. In (Fig. 6c–d) we show the distribution of the overall accuracy across teams and split the value by forecasted weeks ahead and observed category. In the 2024–25 season, models were less accurate when trends were increasing compared to decreasing and stable regimes. Further, across both seasons, within each directional category, large increase and large decrease had lower accuracy than increase or decrease, highlighting the difficulty in predicting large trend shifts.

## 3. Discussion

In this work we present a framework for evaluating categorical trend forecasts of laboratory-confirmed influenza hospital admissions submitted to the CDC FluSight Challenge. These forecasts are meant to communicate the probability of directional change, which could more directly inform many questions public health officials tend to ask: are hospital admissions likely to go up, down, or remain steady, and how confident should we be in these predictions? In contrast, count-based forecasts of weekly influenza hospital admissions require additional context to interpret forecasted future trends. By developing and evaluating forecasts in a categorical framework, we are able to assess whether models can correctly anticipate the direction of change, rather than focusing on how close their predictions are to the eventually observed number of influenza hospital admissions.

We quantified categorical trend forecast performance in three ways: *category match* (whether a model’s highest-probability category matched the observed trend), *skill scores* (which measure performance relative to a reference forecast), and *error characterization* (which describes the type and direction of erroneous forecasts). Category match was high for 1-week-ahead forecasts but declined substantially at longer horizons, particularly during periods of increasing influenza hospital admissions. We also found substantial variation in performance across individual models, and most models outperformed the equal probability reference model (Table 1). Similar to how changes in threshold definitions change the distribution of observed categories (Figure S2), we expect that the threshold definitions could impact observed relative performance across the different categories. Consistent with other infectious disease forecasting challenges^7–9^, ensemble approaches were among the top-performing models on average over the seasons we evaluated. The performance of ensembles was robust to the choice of construction method. Averaging probabilities directly from submitted categorical forecasts (FluSight-ensemble) and converting the standard FluSight quantile ensemble of weekly influenza hospital admission counts into categorical probabilities (FluSight-ens_q_cat) yielded similar Brier skill scores.

In the 2024–25 season, nearly all models performed worse than the reference model (i.e. received negative skill scores) during the two periods of increasing influenza hospital admissions in late December 2024 and late January 2025 (Fig. 4a). In the 2025–26 season, most models performed worse than the reference model during the prolonged decline in influenza hospital admissions later in the season (Fig. 4b), driven by rising influenza B activity and declining influenza A activity. Models were most accurate when predicting stable periods and least accurate when predicting categories of large increases and large decreases (Fig. 6). However, we were unable to characterize early-season performance due to delays in data availability, which prevented the forecasting challenges from starting until mid-November in both seasons. Across nearly all models in both seasons, forecasts systematically underestimated the observed trend and were more likely to predict stable or increases/decreases than large increases or large decreases. These observations mirror performance trends observed for influenza hospital admission count forecasts^9–11^ (Fig. S6). Advances in forecasting methodology are needed to better anticipate and characterize changes in influenza hospital admissions, particularly since these periods of change are likely to be most valuable for informing public health efforts to mitigate influenza morbidity and mortality.

To justify our findings and choices in both reference models and scoring rules, we performed multiple sensitivity analyses. The choice of reference model shapes what a skill score communicates to a forecast user. Here, we compare two models, one that assumes the same probability for all trends and one derived from a flat baseline forecast (FluSight-baseline_cat)^30^. Our evaluation shows that model rankings were relatively consistent across the two approaches. However, individual models showed higher skill scores against the FluSight-baseline_cat, particularly during periods of change, reflecting that most models better anticipate directional shifts than this baseline. Each reference has limitations, the equal-category model is uninformative by design, while the baseline model ignores recent trends. When interpreting skills scores, it is important to understand the context of these references. Future forecasting challenges may benefit from consideration of reference models designed so that skill against the reference reflects practical value for decision-making.

Similarly to the reference model, the choice of the scoring rule shapes how models are penalized. The Brier Score does not account for the fact that the categorical trends are ordered (e.g., predicting an increase when there is a decrease should be considered worse than predicting a decrease when there is a large decrease). The ranked probability score accounts for the order of categories (Sec. 2.2.3). While there are some fluctuations in the pairwise ordering of models, the aggregate skill scores are well correlated across both metrics (Fig. 5b). This suggests that our analysis is robust to the choice of scoring rule. As with the reference model, the appropriate scoring rule depends on the use case, and when direction of change matters, the ranked probability score could be more informative.

Our findings have implications for how categorical trend forecasts could be generated and used operationally. The strong performance of the FluSight-ens_q_cat model, which derives categorical probabilities directly from the traditional count-based FluSight quantile ensemble, suggests that categorical probabilities may be derived directly from ensembled, quantile forecast submissions without impairing forecast performance. This observation of our study is likely a consequence of how teams generated their submissions, each of the submitted models in 2024-25 and 2025-26 computed categorical probabilities from their count-based forecast models: either using underlying forecast trajectories or quantiles generated from their submitted forecast distributions. In future seasons, the traditional count-based, quantile submission could therefore be repurposed to produce categorical forecasts. Categorical communication products could then be tailored to specific needs or jurisdictions and produced for different use cases without adding the burden on participating teams of adding additional forecast targets. There remains an opportunity, however, for models designed specifically to predict the probabilities of change directly, rather than as a derived quantity. This could be particularly valuable for improving forecasts during periods of rapid change, where count-based forecasts often struggle.

A perennial challenge in infectious disease forecasting is how to communicate uncertainty in a way that is accessible to non-experts and supports public health decision-making. Count-based forecasts and prediction intervals have become standard ways of presenting forecasts with uncertainty in collaborative forecasting hubs, but they may not be readily interpretable by broader audiences. By directly communicating the chance that influenza hospital admissions will increase, decrease, or be stable, categorical trend forecasts offer a complementary alternative.

This analysis found there are several opportunities for improving the utility of categorical forecasts. Model development is needed to better anticipate periods of rapid change, where current forecasts struggle most. While the evaluation that we performed here can motivate model development and improvements in performance, other evaluations will likely be needed to build user trust. Communication products built on categorical forecasts, including plain-language summaries of expected upcoming trends, need development and testing with the audiences they are meant to serve. Categorical forecast evaluations, such as those presented here, will play an important role in assessing the reliability and robustness of these forecasts to build user trust in their subsequent communication and support their use to inform public health decision-making.

## 4. Methods

### 4.1 Surveillance data

We use the National Healthcare Safety Network (NHSN) dataset of weekly laboratory-confirmed influenza hospital admissions^31^, restricting our analysis to periods of weekly mandatory reporting (see SI for details on reporting cadence). NHSN data are subject to revision as additional reports are received or corrected. These updates, referred to as backfill, are typically positive because preliminary reports may be incomplete and can affect forecast performance. We characterized backfill by comparing the values observed at the time the forecasts were made with the values reported as of 2026-07-01, which were considered final values for the purpose of this analysis. This limitation is discussed in Sec. S6 of the SI. This study spans two influenza seasons (2024–2025, 2025–2026) and includes 52 U.S. jurisdictions (50 states, the District of Columbia, and Puerto Rico) and national level data. In each season, models submitted weekly categorical trend forecasts of laboratory-confirmed hospital admissions (see FluSight GitHub repositories^13^ for full submission details).

### 4.2 Categorical Definitions

Forecast skill was evaluated against the FluSight rate-trend target, which categorizes the trajectory of weekly confirmed influenza hospital admissions into five ordered classes: *large decrease*, *decrease*, *stable*, *increase*, and *large increase*. The hospital data used are taken from the NHSN^30^, which is updated weekly during the influenza forecasting season. Categories are defined by the change in state-level admission incidence (counts per 100,000 population) between the most recently reported baseline week and the target end date for a given week ahead, *w* (population counts have been updated with the latest U.S. census data for the 2025-2026 season^31^):

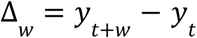

where *t* is the forecast week and *y*_*t*_ is the finalized hospitalization rate for the week *t*. Week pairs with an absolute count difference below 10 admissions are classified as *stable* irrespective of rate change. Forecasted week ahead-specific rate thresholds separating the remaining categories were derived from historical FluSurv-NET and NHSN distributions and are provided in Table 2 (see additional details on construction in Mathis et. al 2025 ^14^). FluSurv-NET provides a long-term population-based surveillance record dating to the 2005-06 season and covering approximately 10% of the U.S. population, whereas NHSN provides nationwide hospitalization surveillance beginning with the 2021-22 influenza season. Forecasts consist of a probability vector across the five categories summing to one. Further detail on the category specifications and corresponding characteristics can be found in SI Sec. S1.

**Table 2:**
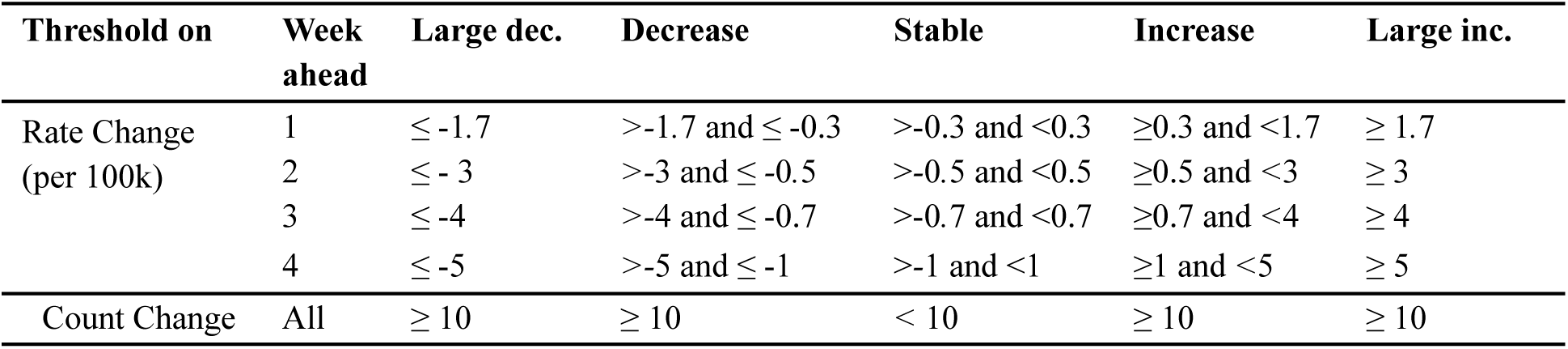
Thresholds for rate changes (per 100k) across horizons in season 2024–25 and 2025–26.

**Table 3:**
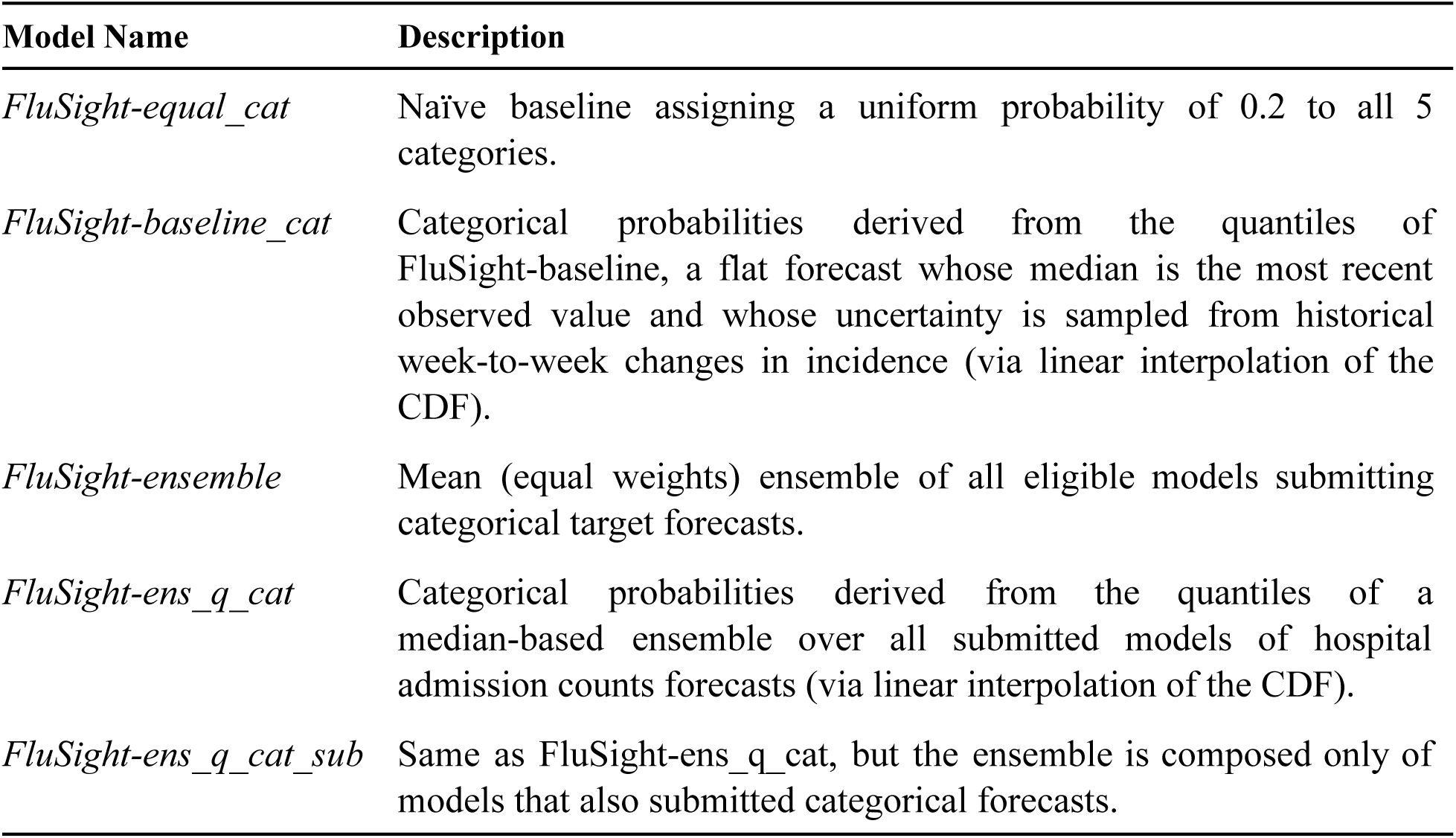
Additional forecast models generated and evaluated alongside individual team models.

| Model Name | Description |
| --- | --- |
| *FluSight-equal_cat* | Naïve baseline assigning a uniform probability of 0.2 to all 5 categories. |
| *FluSight-baseline_cat* | Categorical probabilities derived from the quantiles of FluSight-baseline, a flat forecast whose median is the most recent observed value and whose uncertainty is sampled from historical week-to-week changes in incidence (via linear interpolation of the CDF). |
| *FluSight-ensemble* | Mean (equal weights) ensemble of all eligible models submitting categorical target forecasts. |
| *FluSight-ens_q_cat* | Categorical probabilities derived from the quantiles of a median-based ensemble over all submitted models of hospital admission counts forecasts (via linear interpolation of the CDF). |
| *FluSight-ens_q_cat_sub* | Same as FluSight-ens_q_cat, but the ensemble is composed only of models that also submitted categorical forecasts. |

### 4.3 Forecast model descriptions

We evaluated forecasts submitted during the 2024–25 and 2025–26 influenza seasons, For the 2024–25 season, forecasts from 20 different models were evaluated for reference dates between November 23, 2024, and May 31, 2025, excluding January 25, 2025 (due to data reporting delays), with corresponding target dates from November 23, 2024, to June 21, 2025. Evaluation for the 2025–26 season began on November 22, 2025, and ended on May 23, 2026, and included 16 different teams. In both seasons, forecasts were evaluated only when corresponding ground truth was available. Along with individual models, we generated and evaluated five additional forecast models, two baseline models and three ensemble models defined in Table 3.

### 4.4 Evaluation metrics

We evaluate model performance using the Brier Score^29^ and the Ranked Probability Score (RPS)^33–35^, and explore different approaches to normalizing and aggregating these metrics across jurisdictions and forecast targets. Skill scores are obtained by comparing their performance with naive baseline model predictions (Table 3). We consider Brier Scores as our main metric, due to their relative familiarity, and primarily evaluate forecast performance against the equal probability model, FluSight_equal_cat (see SI Sec. S5).

### Brier Score

Define a numeric index for the *K* ordered categories *C* = {1,…,*K*}. For a given location l, forecast week t, and forecast horizon *a*, let *Y*_*l,t,a*_ denote the observed category and 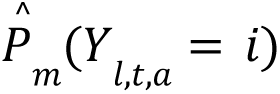 denote the probability assigned by model *m* to category *i*. Then, the Brier Score (BS) measures the squared difference between the forecast probabilities and the observed outcome. *m*￼(*l, t, a*) ￼ is then defined as

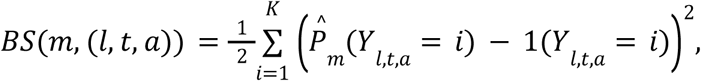

where 1(*Y*_*l,t,a*_ = *i*) denotes the indicator of the event that observed category is *i*.

### RPS

Define cumulative distribution function (CDF) over the category index as:

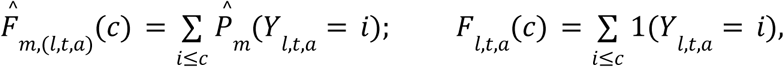

for *c*∈{1,…,*K*}. The Ranked Probability Score (RPS) for model *m* at forecast target (*l, t, a*) is then defined as

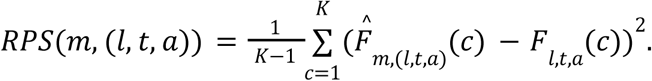

The normalization by 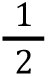 in BS and 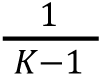 in RPS ensures that the score remains between [0, 1], where 0 indicates a perfect forecast (probability 1 for the true category) and 1 indicates a fully incorrect forecast (probability 1 assigned to incorrect category). In the special case of two categories, the RPS reduces to the BS. The BS is a strictly proper scoring rule for nominal (unordered) categorical outcomes, measuring the mean squared difference between forecast probabilities and the observed category, represented by assigning a value of 1 to the observed category and 0 to all others.^29^ However, when categories have a natural order (e.g., *large decrease*, *decrease*, *stable*, *increase*, or *large increase*), BS does not distinguish whether probability mass placed on incorrect categories was *near* the true category or far from it.

The Ranked Probability Score (RPS) is designed for such *ordinal* settings: it compares cumulative probabilities across ordered categories so that a near miss is penalized less than a far miss.^36^ In particular, forecast assigning all probability to an incorrect category gives identical BS irrespective of their ordinal distance, whereas RPS varies proportionally with this distance. Further illustrative examples are provided in the SI (Sec. S4.2).

### 4.5 Skills for model evaluation

In order to better contextualize the scores, we transform scores for each submitted model *m* into **Brier Skill Score (BSS)** values relative to the reference model *r*. We define skill score for a given location *l*, forecast week *t*, and number of weeks ahead *a* as:

**BSS for target** (*l, t, a*):

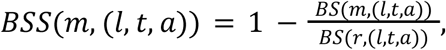

When summarizing BSS across multiple targets, we use the same aggregation method used to prepare some “relative WIS” skill evaluations used in quantile forecast evaluation^8^:

### Overall aggregate BSS

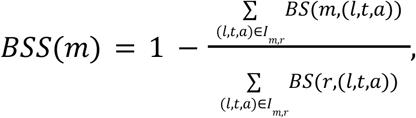

where *I*_*m,r*_ is the intersection of the availability of *m* and of *r*, that is, the set of (l, t, a) prediction targets for which both *m* and *r* submitted predictions.

When breaking down model performance by week ahead or by eventually-observed rate-trend category, we use the following definition for BSS on each subset of targets:

### Aggregate BSS on a subset of targets ***S***

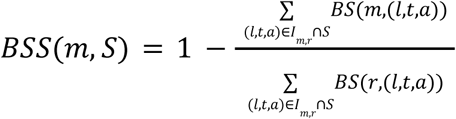

Using the main reference model FluSight-equal_cat, individual-target and aggregate BSS values can range from-1.5 to 1, with higher being better, and 0 indicating no improvement over the reference model. Sec. S5 details other properties of skill scores. For sensitivity analysis, we define a Ranked Probability Skill Score (RPSS) analogously, by replacing BSS and BS with RPSS and RPS.

### 4.6 Probability weighted, multi-category accuracy

We define a probability weighted, multi-category accuracy metric to highlight the performance of individual modeling teams. Let *Y*_*l,t,a*_ denote the observed categorical outcome for location *l*, forecast week *t*, and number of weeks ahead *a*, and let 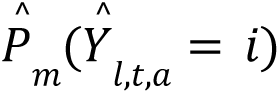 be the probability assigned by model *m* to category *i* for that instance. The column-normalized probabilistic confusion matrix entry for forecast category *i* and observed category *j* is defined as

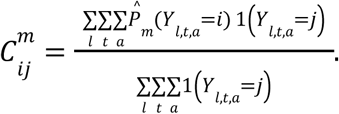

Thus, *C_ij_^m^* represents the average probability mass assigned to forecast category *i* across all forecasts for which the observed FluSight category was *j*, and each column of *C*^*m*^ sums to one. We further evaluate the performance of individual modeling teams using a conditional accuracy metric. For a given categorical trend *j*, the conditional accuracy is defined as the diagonal element of the probabilistic confusion matrix, *C_ij_^m^*, and measures the average probability mass that model *m* assigns to the correct category conditional on that category being observed.

### Disclaimer

The contents of this manuscript are solely the responsibility of the authors and do not necessarily represent the official views of the Centers for Disease Control and Prevention, the U.S. National Institutes of Health, the U.S. Department of Health and Human Services, the Council of State and Territorial Epidemiologists, the National Science Foundation, or other funding organizations. Any use of trade, firm, or product names is for descriptive purposes only and does not imply endorsement by the U.S. Government.

## Contributions

J.T.D., G.K., A.H., M.B.N., S.Venkatramanan, L.B., S.Mathis, and R.K.B. conceived and designed the study, curated the data, performed the analysis, and wrote the original draft. J.T.D., G.K., A.H., M.B.N., S.Venkatramanan, L.B., S.Mathis, M.A., M.L., A.G.K., P.C.V., S.Mhade, D.Weber, D.S., N.D., D.J.M., T.Y., R.Z.T., J.S., R.Y., S.Pei, C.B., A.W., L.S., E.R., S.W., J.N., W.T.R., L.Mullany, R.Posner, A.M., Y.T.L., W.S.H., A.S., A.A.G., A.D., B.J.F., E.F.M., J.Lee, J.R.M., K.A.G., M.Baltz, O.B., Y.M.B.T., Y.C., C.J.G., M.Santillana, C.D., J.Lu, S.Y., F.L., L.C., A.G.M., A.U., N.G., M.Chinazzi, M.U., N.M., R.LeWinter, S.Venturini, S.F., A.V., S.J.F., E.S., M.Salcedo, R.Prabhakar, B.K.M.C., A.P., C.V., J.T., V.P.N., A.B., D.Williams, G.C.G., L.Meyers, E.T., C.V.B., R.Loster, B.B.L., M.Cojocaru, P.Y., A.R., R.Li, S.Potnis, N.G.R., T.R., J.Lemaitre, A.A., B.L., M.M., N.C.M., S.D.T., N.R., Y.S., and R.K.B. submitted forecast data for the analysis. All authors contributed to the review and editing of the manuscript.

## Data Availability

The forecast data for each model are publicly accessible from the FluSight Forecast Hub GitHub repository(https://github.com/cdcepi/FluSight-forecast-hub^13^; https://doi.org/10.5281/zenodo.22101290). The target data are also available as weekly counts for each jurisdiction from HHS^31^.

## Code Availability

The code used to generate all figures and tables in the manuscript will be made available in a public repository (https://github.com/cdcepi/FluSight-manuscripts).

## Data Availability

The forecast data for each model are publicly accessible from the FluSight Forecast Hub GitHub repository(https://github.com/cdcepi/FluSight-forecast-hub; https://doi.org/10.5281/zenodo.22101290). The target data are also available as weekly counts for each jurisdiction from HHS

https://doi.org/10.5281/zenodo.22101290

## Acknowledgements

J.T.D., M.A., M.L., A.G.K., P.C.V., S.Mhade, C.B., W.T.R., L.Mullany, M.Santillana, A.U., M.Chinazzi, M.U., N.M., R.LeWinter, S.Venturini, A.V., S.J.F., E.S., M.Salcedo, R.Prabhakar, B.K.M.C., J.Lemaitre, and M.J. acknowledge the support of the Insight Net cooperative agreement CDC-RFA-FT-23-0069 from the CDC’s Center for Forecasting and Outbreak Analytics. G.K., M.B.N., S.Venkatramanan, S.J.F., E.S., M.Salcedo, R.Prabhakar, B.K.M.C., J.T., G.C.G., L.Meyers, A.A., B.L., and M.M. acknowledge support from CSTE/CDC cooperative agreement NU38OT000297. J.T.D., G.K., S.Venkatramanan, J.S., C.B., V.P.N., A.B., D.Williams, A.A., B.L., M.M., and S.D.T. acknowledge support from CSTE/CDC cooperative agreement NU38PW000005. L.B., D.Weber, D.S., and N.D. acknowledge support from the Centers for Disease Control and Prevention under award number U01IP001121. L.B., D.Weber, D.S., N.D., and D.J.M. acknowledge support from the United States of America Department of Health and Human Services and Centers for Disease Control and Prevention under contract number 75D30123C15907. L.B. and D.J.M. acknowledge support from the Council of State and Territorial Epidemiologists under a subaward of NU38OT000297. D.J.M. acknowledges the support of the Natural Sciences and Engineering Research Council of Canada (NSERC), ALLRP 581756-23. Cette recherche a été financée par le Conseil de recherches en sciences naturelles et en génie du Canada (CRSNG), ALLRP 581756-23. L.S., E.R., N.G.R., and T.R. acknowledge support from the National Institute of General Medical Sciences (R35GM119582) and the US Centers for Disease Control and Prevention (U01IP001122). G.K., S.Venkatramanan, A.A., B.L., M.M., and N.C.M. acknowledge support from NSF Expeditions in Computing grant CCF-1918656. G.K., S.Venkatramanan, A.A., B.L., and M.M. acknowledge support from NIGMS R24GM153920 (MIDAS Coordination Center) and CDC-PGCoE grant 6NU50CK000555-03-01. J.S. acknowledges support from National Institute of Allergy and Infectious Diseases grant R01AI163023. S.Pei acknowledges support from National Institute of General Medical Sciences grant R35GM156799 and National Science Foundation grant DMS-2229605. R.Posner and W.S.H. acknowledge support from National Institutes of Health / National Institute of General Medical Sciences grant R01GM111510. J.Lemaitre acknowledges support from the National Institutes of Health (NIH, award 5R01AI102939). N.R. and Y.S. acknowledge support from National Science Foundation grant DBI-2412389.

