## Supplementary Information for "Multi-season evaluation and analysis of categorical trend forecasts of influenza hospital admissions in the United States"

### Supplementary Information: Multi-season evaluation of probabilistic trend forecasts of influenza hospitalizations in the United States

#### 1. FluSight Forecast Hub

The CDC FluSight Forecasting Challenge is a collaborative forecasting initiative that solicits weekly probabilistic forecasts of influenza activity from modeling teams across academia, government, and the private sector. Since the 2021–2022 season, the challenge has focused on forecasts of laboratory-confirmed influenza hospital admissions reported through NHSN[1], with teams submitting quantile-based probabilistic forecasts of weekly admission counts for each US jurisdiction and nationally at 1-4 weeks ahead.

FluSight introduced a categorical trend target in which teams submit probability distributions over five ordered categories (*large decrease, decrease, stable, increase, large increase*) describing the expected change in influenza hospital admission rates. Categories are determined by the change in the jurisdiction-level hospital admission rate (per 100,000 population) between the reference week and the target week at each forecast horizon. Week pairs with an absolute count difference below 10 admissions are classified as *stable* regardless of rate change; the remaining categories are separated by week-ahead specific rate thresholds derived from historical FluSurv-NET and NHSN week over week distributions[2] (Table S1).

***Table S1: Thresholds for rate (per 100k) and count changes across horizons (2024-25, and 2025–26 seasons).***

| Threshold on | Week ahead | Large dec. | Decrease | Stable | Increase | Large inc. |
|---|---|---|---|---|---|---|
| Rate Change (per 100k) | 1 | ≤ -1.7 | >-1.7 and ≤ -0.3 | >-0.3 and <0.3 | ≥0.3 and <1.7 | ≥ 1.7 |
| | 2 | ≤ - 3 | >-3 and ≤ -0.5 | >-0.5 and <0.5 | ≥0.5 and <3 | ≥ 3 |
| | 3 | ≤ -4 | >-4 and ≤ -0.7 | >-0.7 and <0.7 | ≥0.7 and <4 | ≥ 4 |
| | 4 | ≤ -5 | >-5 and ≤ -1 | >-1 and <1 | ≥1 and <5 | ≥ 5 |
| Count Change | All | ≥ 10 | ≥ 10 | < 10 | ≥ 10 | ≥ 10 |

#### *1.1 Observed categorical trends*

Each season produces a different distribution of observed categories (Table S2). In 2024–25, the distribution was much more spread out: stable weeks fell to 29–37% across horizons (32.0% overall), decreases (decrease plus large decrease) made up about 39.1% of targets, and increases (increase plus large increase) about 28.9%. The 2025–26 season shows a similar shape to 2024–25 but with more weight on stable weeks: stable weeks ranged from 41–47% across horizons (43.4% overall), decreases made up about 33.8% of targets, and increases about 22.8%. Within each season, the distribution is fairly consistent across horizons, though the share of stable targets falls. Fig S1 shows the observed categorical trends across all jurisdictions and nationally for the 2024–25 and 2025–26 seasons at each forecast horizon. The observed categories for the 1-4 week ahead targets are visualized by the date the forecast was generated (all called the reference date) not the exact forecast date.

***Table S2: Target category distribution for the 2024–25 and 2025–26 seasons by forecast horizon.*** *Each row reports the number of reference weeks, jurisdictions, and total target instances scored, followed by the count (percentage) in each rate-change category. The "All" row aggregates across horizons within a season.*

| Season | Week ahead | Weeks | Jurisdictions | Total | Large dec. | Decrease | Stable | Increase | Large inc. |
|---|---|---|---|---|---|---|---|---|---|
| 2024/25 | 1 | 27 | 53 | 1431 | 239 (16.7%) | 279 (19.5%) | 534 (37.3%) | 160 (11.2%) | 219 (15.3%) |
| | 2 | 27 | 53 | 1431 | 249 (17.4%) | 318 (22.2%) | 465 (32.5%) | 189 (13.2%) | 210 (14.7%) |
| | 3 | 27 | 53 | 1431 | 259 (18.1%) | 322 (22.5%) | 418 (29.2%) | 204 (14.3%) | 228 (15.9%) |
| | 4 | 27 | 53 | 1431 | 245 (17.1%) | 325 (22.7%) | 417 (29.1%) | 194 (13.6%) | 250 (17.5%) |
| | **All** | 31 | 53 | 5724 | 992 (17.3%) | 1244 (21.7%) | 1834 (32.0%) | 747 (13.1%) | 907 (15.8%) |
| 2025/26 | 1 | 27 | 53 | 1431 | 141 (9.9%) | 279 (19.5%) | 675 (47.2%) | 187 (13.1%) | 149 (10.4%) |
| | 2 | 27 | 53 | 1431 | 144 (10.1%) | 334 (23.3%) | 622 (43.5%) | 189 (13.2%) | 142 (9.9%) |
| | 3 | 27 | 53 | 1431 | 136 (9.5%) | 379 (26.5%) | 582 (40.7%) | 180 (12.6%) | 154 (10.8%) |
| | 4 | 27 | 53 | 1431 | 122 (8.5%) | 401 (28.0%) | 606 (42.3%) | 141 (9.9%) | 161 (11.3%) |
| | **All** | 30 | 53 | 5724 | 543 (9.5%) | 1393 (24.3%) | 2485 (43.4%) | 697 (12.2%) | 606 (10.6%) |

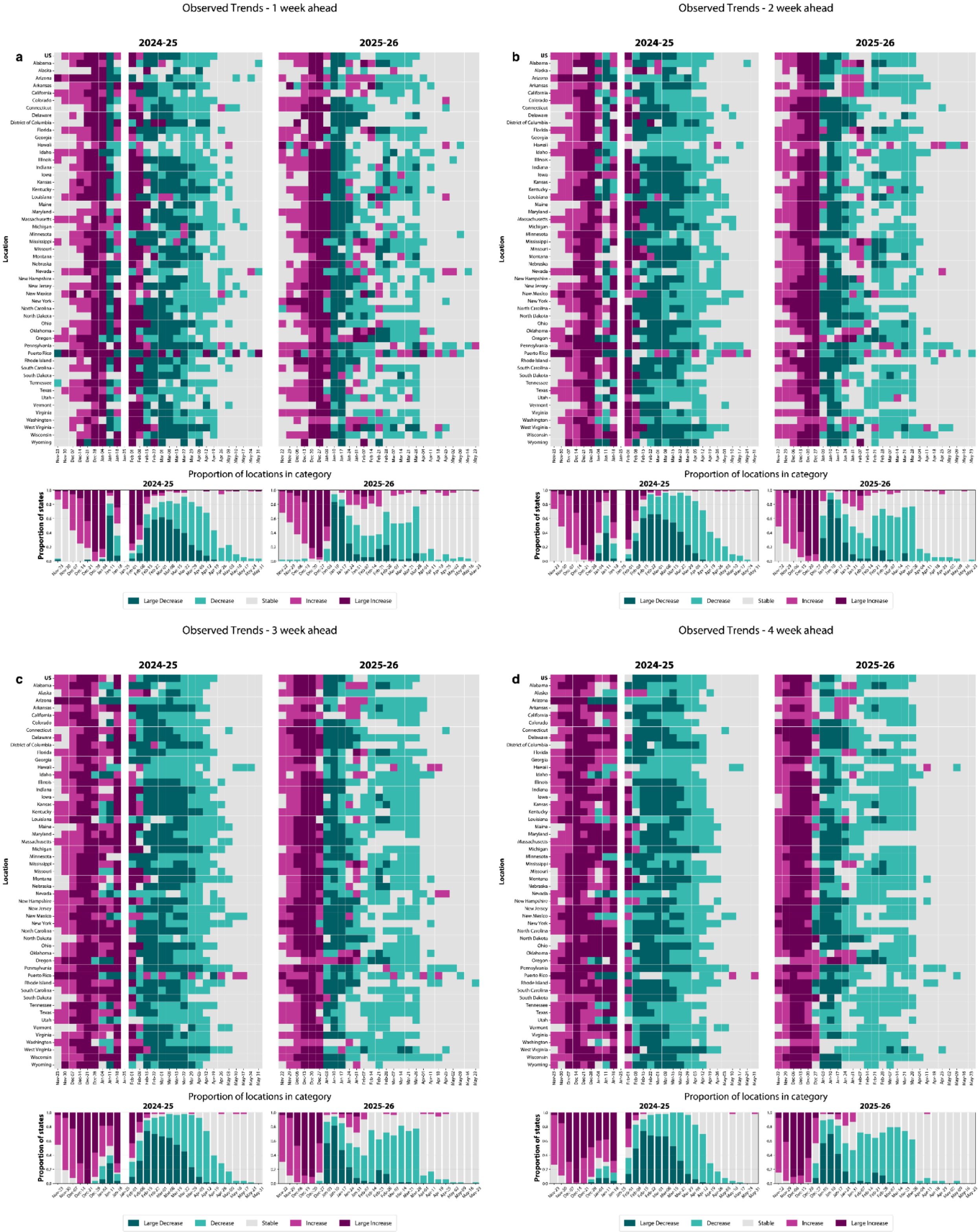

***Figure S1: The observed trends of the 2024–25 and 2025–26 seasons for 1–4 weeks ahead (a-d).*** *Within each subfigure, the top row of heatmaps shows the observed change of each jurisdiction for each week in the two seasons. The bottom row shows the proportion of jurisdictions within each category during a given week.*

#### 1.2 *Impact of category definitions*

The choice of threshold value for the trends is important and can affect forecast performance as well as the utility of the trend forecasts themselves. Although the current text focuses on the performance of categorical forecasts in 2024–25 and 2025–26, categorical predictions were also submitted during the 2023–24 season. While a full analysis of that season is outside the scope of this work, the definitions of the thresholds used to define the categories were different in 2023–24 compared to 2024–25 and 2025–26 (Table S3). The stable-category threshold was wider at every horizon in 2023–24 (e.g., $< 1/100$k vs. $< 0.3/100$k at 1-week ahead), with this gap generally increasing at longer horizons. In contrast, the threshold separating large increase/decrease from moderate change was only modestly wider in 2023–24 at 1-week ahead ( $\geq 2/100$k vs. $\geq 1.7/100$k) and was identical to the 2024–25/2025–26 thresholds at 2- through 4-week horizons. We use the 2023–24 threshold definitions as an example to compare how observed trend classifications would have changed under a different set of definitions. As a result, more jurisdiction-weeks were classified as stable under the 2023–24 definitions than would have been under the later, narrower thresholds (Fig. S2). It remains an open question which framing would be most useful for users and likely depends on the use case. For example, decisions about increasing hospital staffing might be better informed by definitions that require larger differences. However, detecting early increases in influenza hospital admissions may be better suited to lower thresholds for change.

***Table S3: Thresholds for rate and count changes across horizons for the 2023–24 season.***

| Threshold on | Week ahead | Large dec. | Decrease | Stable | Increase | Large inc. |
|---|---|---|---|---|---|---|
| Rate Change (per 100k) | 1 | ≤ -2 | >-2 and ≤ -1 | >-1 and <1 | ≥1 and <2 | ≥ 2 |
| | 2 | ≤ -3 | >-3 and ≤ -1 | >-1. and <1 | ≥1 and <3 | ≥ 3 |
| | 3 | ≤ -4 | >-4 and ≤ -2 | >-2 and <2 | ≥2 and <4 | ≥ 4 |
| | 4 | ≤ -5 | >-5 and ≤ -2.5 | >-2.5 and <2.5 | ≥2.5 and <5 | ≥ 5 |
| Count Change | All | ≥ 10 | ≥ 10 | < 10 | ≥ 10 | ≥ 10 |

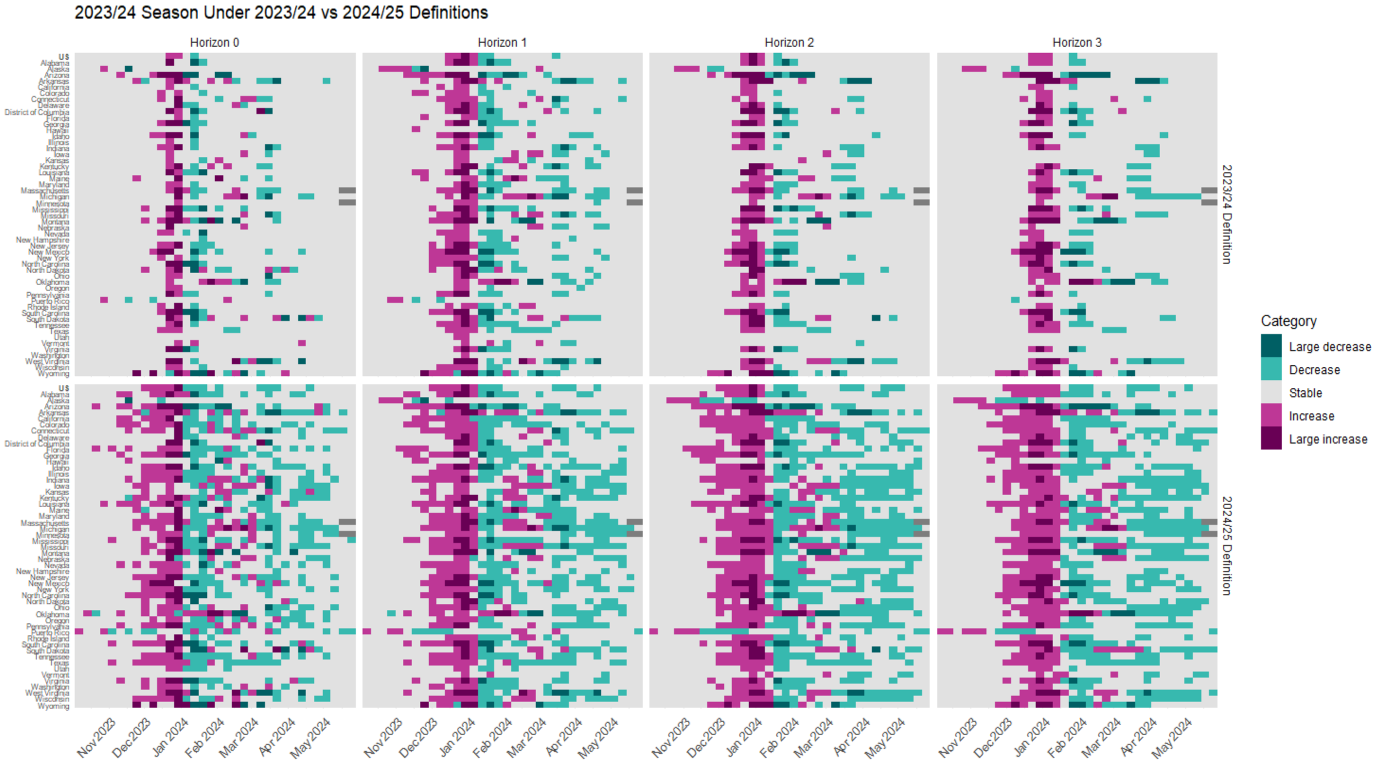

***Figure S2: The observed categorical trends during the 2023–24 season using the 2023–24 season thresholds versus the 2024–25 and 2025–26 thresholds for each week ahead.*** *Each panel shows the observed change of each jurisdiction over each week during the season for the designated category definition.*

#### *2*. Model inclusion criteria for evaluation

The date each forecast is submitted corresponds to a *reference date*, the Saturday ending the week the forecast was submitted (one week after the most recent surveillance data). For the 2024–25 season, the evaluation covered reference dates from November 23, 2024 to May 31, 2025. For the 2025–26 season, we evaluated forecasts starting on November 22, 2025 to May 23, 2026. Forecast submissions were limited to combinations of location, reference date, and horizon for which both a *FluSight-ensemble* forecast and the corresponding observed ground truth were available. This approach is used to ensure that each target for evaluation corresponds to an available ground truth observation. This resulted in 5724 forecasts for both the 2024–25 and 2025–26 seasons. Under these criteria, for the 2024–25 and 2025-26 seasons, across locations, there were 27 weekly targets for all forecast horizons (see Table S4).

Models were eligible if they submitted forecasts for at least 75% of the location–reference date–week-ahead combinations covered by the *FluSight-ensemble*. Using these criteria, 15 models from 2024–2025 and 11 models from 2025–26 met the requirements and were included in the evaluation (excluding the FluSight reference and ensemble models). Detailed counts of the number of forecast submissions and the thresholds for each season are provided in Table S5.

***Table S4: Number of FluSight-ensemble targets per week-ahead, per jurisdiction for all three influenza seasons.***

| Season | 1-wk ahead | 2-wk ahead | 3-wk ahead | 4-wk ahead | Notes |
|---|---|---|---|---|---|
| 2024–25 | 27 | 27 | 27 | 27 | All 53 jurisdictions |
| 2025–26 | 27 | 27 | 27 | 27 | All 53 jurisdictions |

***Table S5. Model inclusion based on forecast frequency threshold for the 2024–25 and 2025–26 seasons.*** *Percentages in brackets are relative to FluSight-ensemble forecasts. An em dash indicates the model did not submit forecasts that season. Note: even though models may have the same name across seasons it is not a given that the modeling team used the same methodology.*

| **Model ID** | **2024–25** | **2025–26** |
|---|---|---|
| CEPH-Rtrend_fluH | 5704 (99.7%) | 5724 (100.0%) |
| CU-ensemble | 5512 (96.3%) | 5512 (96.3%) |
| FluSight-baseline_cat | 5724 (100.0%) | 5724 (100.0%) |
| FluSight-ens_q_cat | 5724 (100.0%) | 5724 (100.0%) |
| FluSight-ens_q_cat_sub | 5724 (100.0%) | 5724 (100.0%) |
| FluSight-ensemble | 5724 (100.0%) | 5724 (100.0%) |
| FluSight-equal_cat | 5724 (100.0%) | 5724 (100.0%) |
| JHUAPL-DMD | 5088 (88.9%) | — |
| MIGHTE-Joint | 4876 (85.2%) | — |
| MIGHTE-Nsemble | 5300 (92.6%) | — |
| MOBS-EpyStrain_Flu | — | 4908 (85.7%) |
| MOBS-GLEAM_FLUH | 4996 (87.3%) | — |
| MOBS-GLEAM_RL_FLUH | — | 5196 (90.8%) |
| NIH-Flu_ARIMA | 5688 (99.4%) | 4976 (86.9%) |
| PSI-PROF | 5724 (100.0%) | 5724 (100.0%) |
| PSI-PROF_beta | 5300 (92.6%) | — |
| PSI-PROF_MOA | — | 5724 (100.0%) |
| SigSci-BECAM | 5524 (96.5%) | — |
| SigSci-TSENS | 5288 (92.4%) | 4644 (81.1%) |
| UGA_flucast-Copycat | 5076 (88.7%) | 5724 (100.0%) |
| UMass-flusion | 5280 (92.2%) | — |
| UMass-trends_ensemble | 5724 (100.0%) | 5724 (100.0%) |
| UVAFluX-Ensemble | 5724 (100.0%) | 5724 (100.0%) |

### 3. Extended performance analysis of the 2024-25 and 2025-26 seasons

#### *3.1 Categorical trend agreement for 2-4 week ahead forecasts*

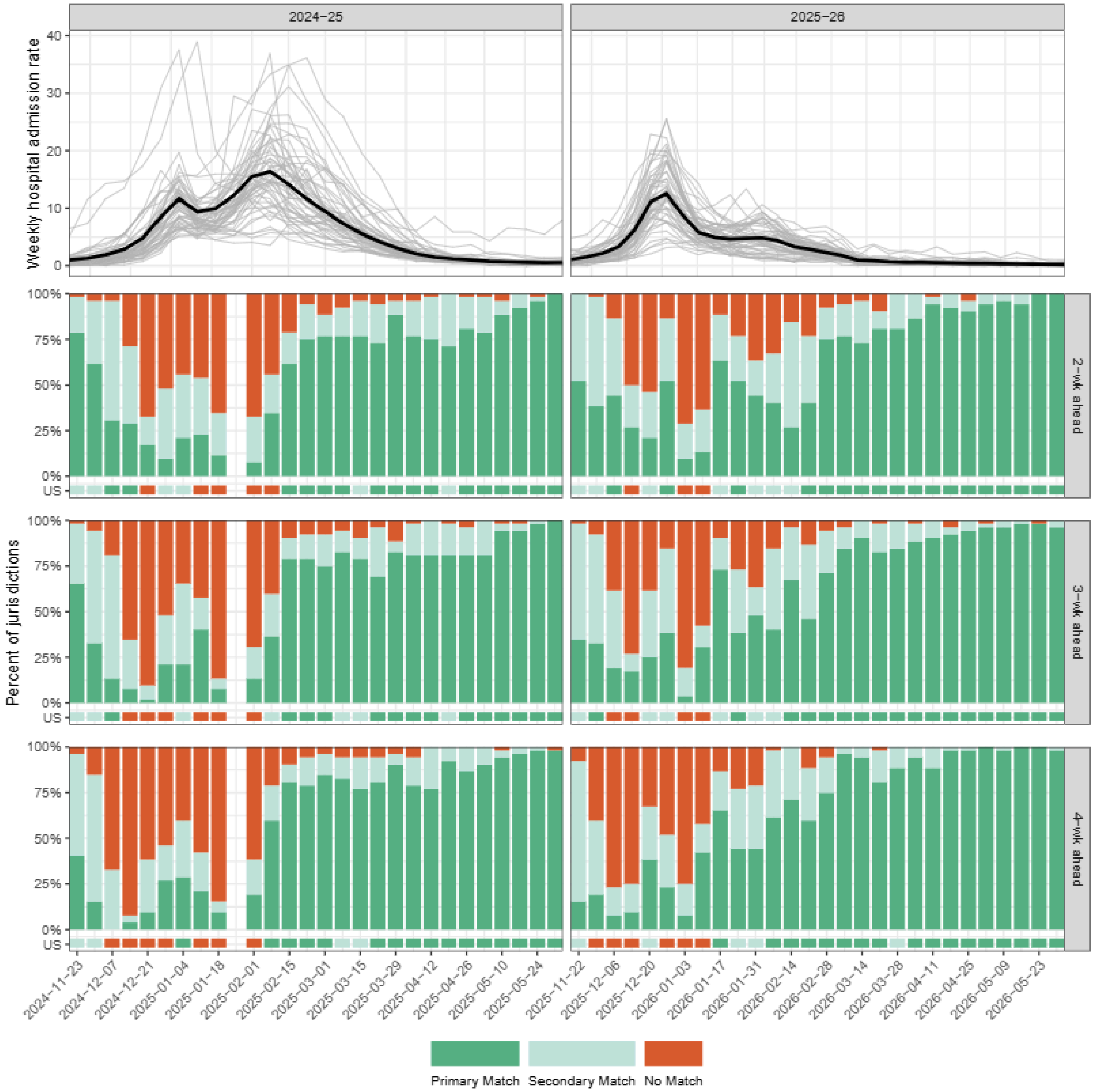

***Figure S3: Observed weekly laboratory-confirmed influenza hospital admissions and corresponding agreement of observed categorical trends with forecasted categorical trends.*** *The top row shows the Epidemic curves for the observed laboratory-confirmed influenza hospital admission rates in the 2024–25 and 2025–26 seasons. Each line represents a single jurisdiction. The following rows show the proportion of jurisdictions within each week that the categorical ensemble (FluSight-ensemble) correctly or incorrectly predicted the observed two to four-week ahead trend category. The Primary match label indicates that the categorical ensemble assigned the highest probability to the observed category. Secondary match indicates that the categorical ensemble assigned the second highest probability to the observed category. Predictions are considered no match if neither the first nor second highest assigned probabilities are observed.*

***Table S6: Agreement of forecasted and observed categorical trends.*** *For each model and season, the table reports the percentage of jurisdiction–week combinations (including the U.S.) for which the forecasted categorical trend aligned with the observed trend for the 1-week-ahead horizon. A primary match indicates that the model assigned the highest probability to the observed category. A secondary match indicates that the model assigned the second-highest probability to the observed category. A forecast is classified as no match if the observed category was neither the highest- nor second-highest-probability category in the forecast.*

| Model | Primary Match (%) | Secondary Match (%) | No Match (%) |
|---|---:|---:|---:|
| **2024–25 Season** | | | |
| FluSight-ensemble | 60.4 | 22.5 | 17.1 |
| PSI-PROF | 60.2 | 20.6 | 19.1 |
| FluSight-ens_q_cat | 59.2 | 22.2 | 18.6 |
| UMass-flusion | 59.1 | 22.5 | 18.4 |
| NIH-Flu_ARIMA | 58.2 | 21.7 | 20.0 |
| FluSight-ens_q_cat_sub | 58.0 | 23.5 | 18.4 |
| PSI-PROF_beta | 57.3 | 22.3 | 20.5 |
| MOBS-GLEAM_FLUH | 55.4 | 24.8 | 19.8 |
| MIGHTE-Nsemble | 54.3 | 19.3 | 26.3 |
| CEPH-Rtrend_fluH | 54.2 | 22.4 | 23.4 |
| MIGHTE-Joint | 53.9 | 21.2 | 24.9 |
| UMass-trends_ensemble | 52.5 | 22.1 | 25.4 |
| CU-ensemble | 51.4 | 25.0 | 23.7 |
| UVAFluX-Ensemble | 47.6 | 26.1 | 26.3 |
| SigSci-TSENS | 46.5 | 24.1 | 29.4 |
| UGA_flucast-Copycat | 41.4 | 24.7 | 33.9 |
| FluSight-baseline_cat | 37.3 | 21.2 | 41.5 |
| SigSci-BECAM | 37.0 | 20.9 | 42.0 |
| JHUAPL-DMD | 34.9 | 13.5 | 51.6 |
| **2025–26 Season** | | | |
| FluSight-ens_q_cat | 66.7 | 19.0 | 14.3 |
| CEPH-Rtrend_fluH | 63.9 | 21.1 | 15.0 |
| FluSight-ensemble | 63.3 | 21.2 | 15.5 |
| FluSight-ens_q_cat_sub | 62.5 | 22.0 | 15.4 |
| MOBS-EpyStrain_Flu | 61.0 | 20.5 | 18.5 |
| MOBS-GLEAM_RL_FLUH | 60.8 | 21.2 | 18.0 |
| NIH-Flu_ARIMA | 58.7 | 23.4 | 17.9 |
| PSI-PROF_MOA | 57.2 | 22.3 | 20.5 |
| PSI-PROF | 56.5 | 22.0 | 21.5 |
| UMass-trends_ensemble | 53.6 | 22.4 | 24.0 |
| UVAFluX-Ensemble | 53.2 | 24.2 | 22.6 |
| UGA_flucast-Copycat | 52.8 | 24.4 | 22.9 |
| CU-ensemble | 51.7 | 26.1 | 22.2 |
| SigSci-TSENS | 50.8 | 22.8 | 26.4 |
| FluSight-baseline_cat | 47.2 | 20.2 | 32.6 |

#### *3.2 FluSight-ensemble model performance across jurisdictions*

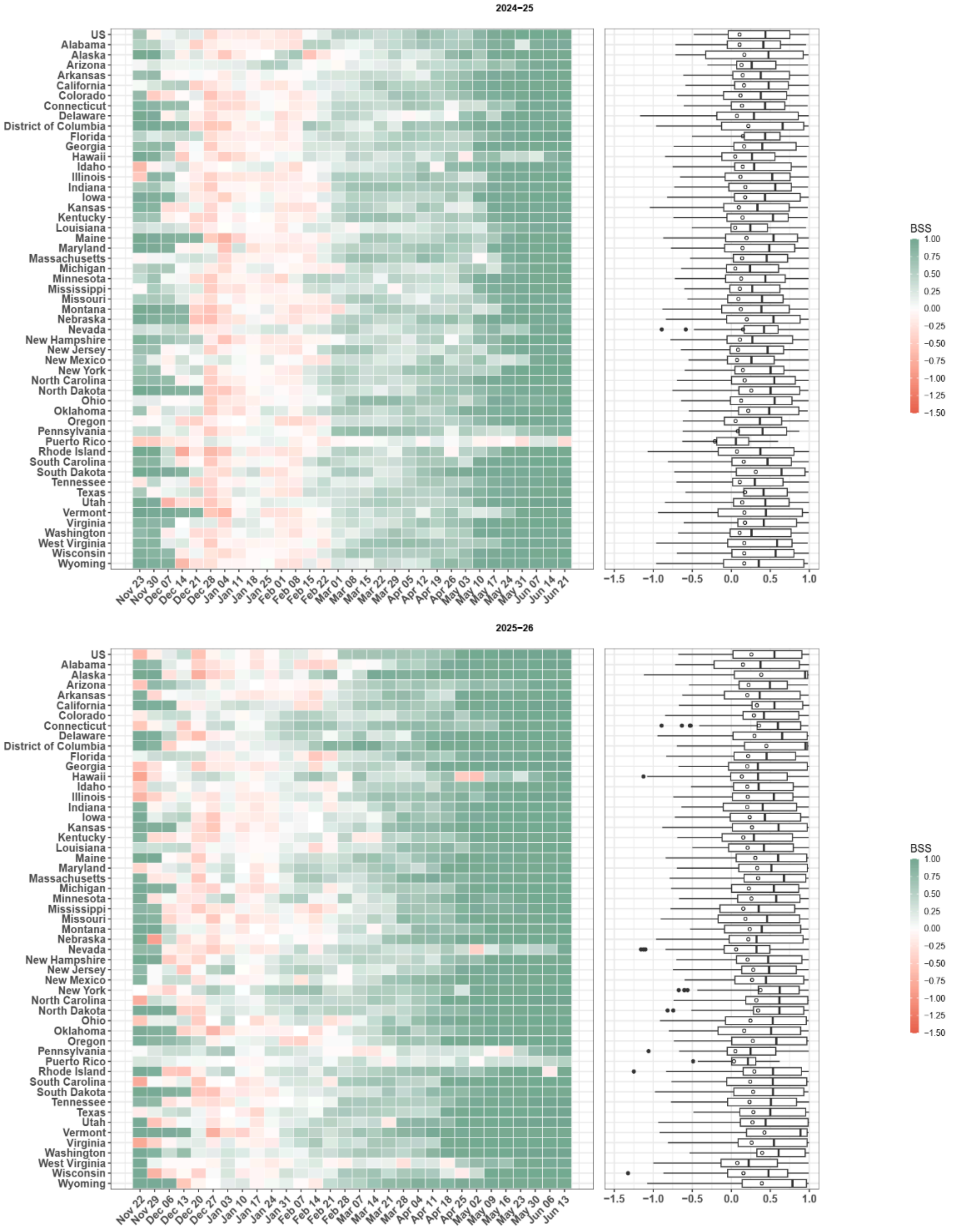

***Figure S4: Spatial variations in the performance of FluSight-ensemble during 2024–25 and 2025–26 influenza seasons.*** *The heatmap shows the mean Brier Skill Score (BSS) for each location, averaged across all 1- to 4-week ahead forecasts for each week. The boxplots summarize the distribution of BSS across 1-4 week-ahead forecast targets. Circles indicate the mean BSS, and the solid vertical lines represent the median BSS.*

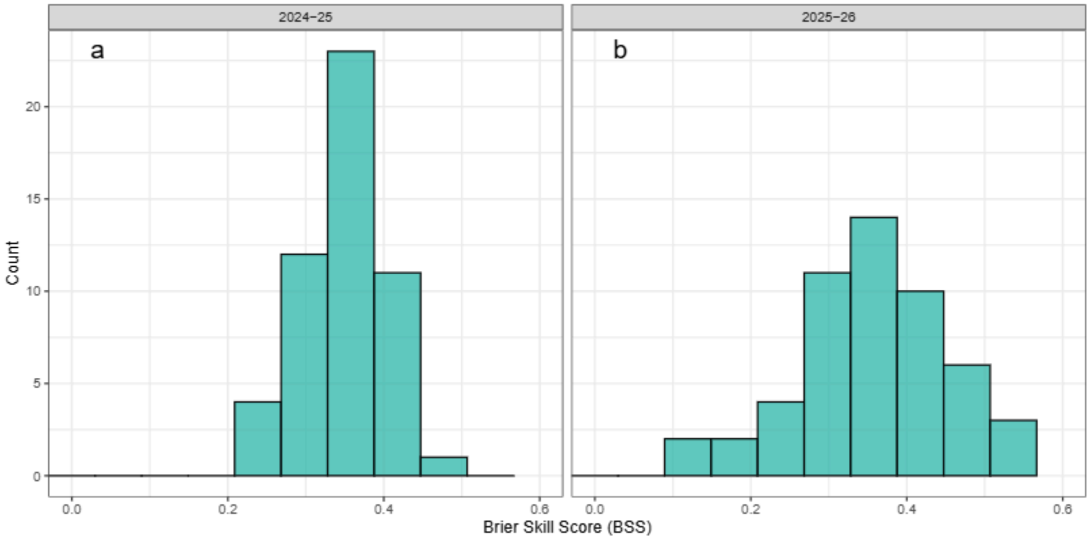

***Figure S5: A histogram representation of the overall Brier Skill Score (BSS) for the FluSight-ensemble model for each location during 2024–25 and 2025–26 seasons (left and right panels).***

#### *3.3 Comparison of count-based forecasts to categorical forecasts*

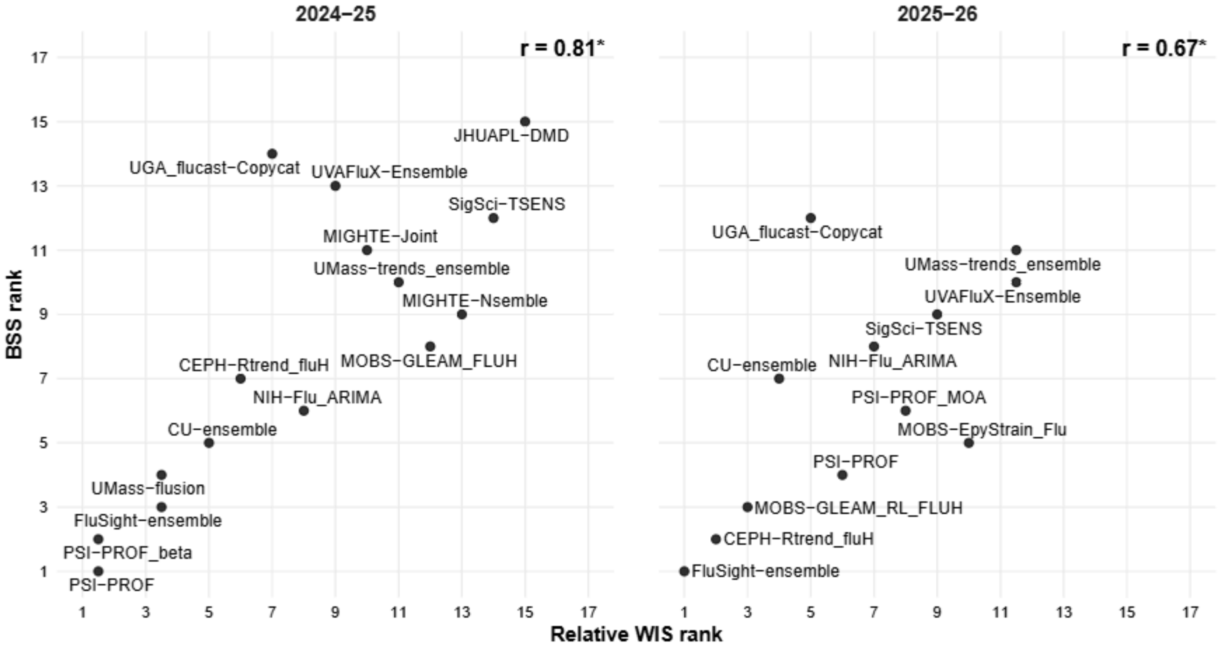

***Figure S6: Rank correlation between categorical trend forecast performance (BSS) and count-based quantile forecast performance (per capita relative WIS) for the 2024–25 (left) and 2025–26 (right) seasons****. Each point represents a model's rank by aggregate BSS plotted against its rank by relative WIS. Lower ranks indicate better performance on each metric. Spearman correlation coefficients are shown for each season (r = 0.81, 2024–25; r = 0.67, 2025–26; both* $p < 0.05$*).*

##### *3.4 Probability-weighted confusion matrices and directional bias*

In the main text we highlighted three modeling teams using confusion matrices with trend categories aggregated into increase, stable, and decrease. Here we present the full probability weighted, multi-category confusion matrices for all teams and both seasons across all five trend categories (Fig. S7, S8). As described in the main text, each cell represents the normalized probability mass forecasted for category $j$ given an observed category $i$, aggregated across all locations, horizons, and forecast weeks. Each column (observed category) is normalized to sum to one, so cell $(i, j)$ is the probability the forecast assigned to category $j$ conditional on observing category $i$, averaged across locations, horizons, and forecast weeks. The center diagonal, from the upper left to lower right, shows the probabilities that the forecasted category correctly matched the observed category. Cells under the diagonal refer to instances where the model overestimates the forecast trend (e.g., a stable or increasing trend was predicted when a decrease was observed). Cells above the diagonal, on the other hand, correspond with underestimates of the forecast trend (e.g., a stable or decreasing trend was predicted when an increase was observed). By taking the difference between the sum of probabilities associated to the cells below and above the diagonals we can define a metric to calculate the directional bias. This metric can be used to assess the symmetry of the forecast errors. We first sum the probabilities where the forecasted categorical trend was “lower” than the observed category (cells above the diagonal, $U$) and the probabilities where the forecasted categorical trend was “higher” than the observed category (cells below the diagonal, $L$). We define the directional bias as $(L - U)/(L + U)$. It ranges from $-1$ (all off-diagonal mass underestimates the trend) to $+1$ (all overestimates), with $0$ indicating symmetric errors. An important note is that the conditional normalization (normalizing the columns to sum to one) causes each observed category to contribute equally to the directional-bias metric regardless of how frequently it occurred.

Models show a systematic underestimation of the trend, which is consistent with the difficulty models have in anticipating increases, which produces a tendency to assign probability mass to milder categories than the one ultimately observed. This pattern also parallels findings from quantile-based forecast evaluations, where stable periods are consistently the easiest to predict and rapid changes the hardest.

We additionally extract the conditional accuracy by category, looking at the diagonal values where the observed and forecasted categories matched. We measured the correlation between these values and BSS and found a strong, consistent positive relationship across both seasons and all forecasted weeks ahead (Fig. S9), with $R^2$ ranging from 0.63 to 0.71.

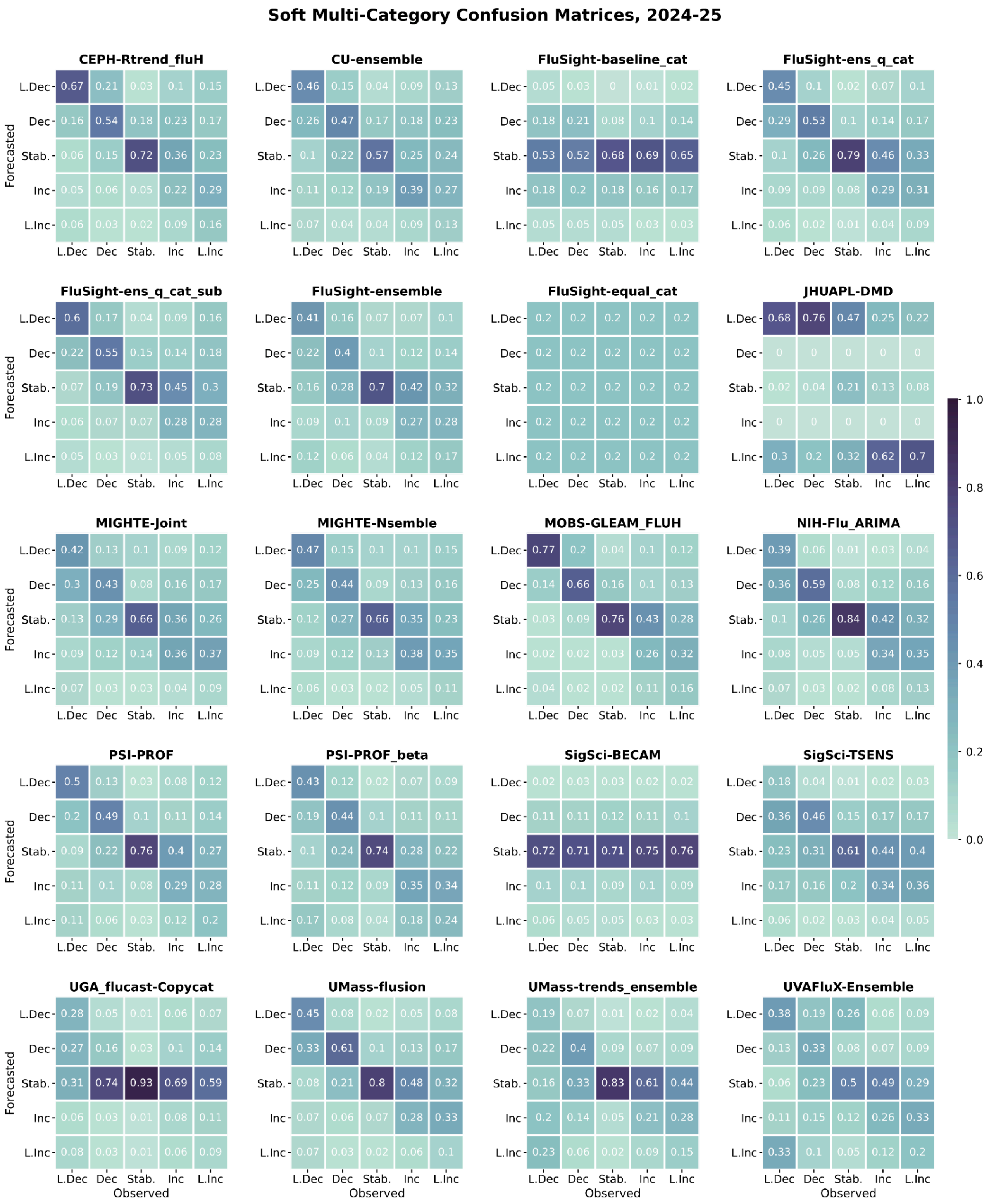

*Figure S7: Confusion Matrices for all teams during the 2024–25 flu season.*

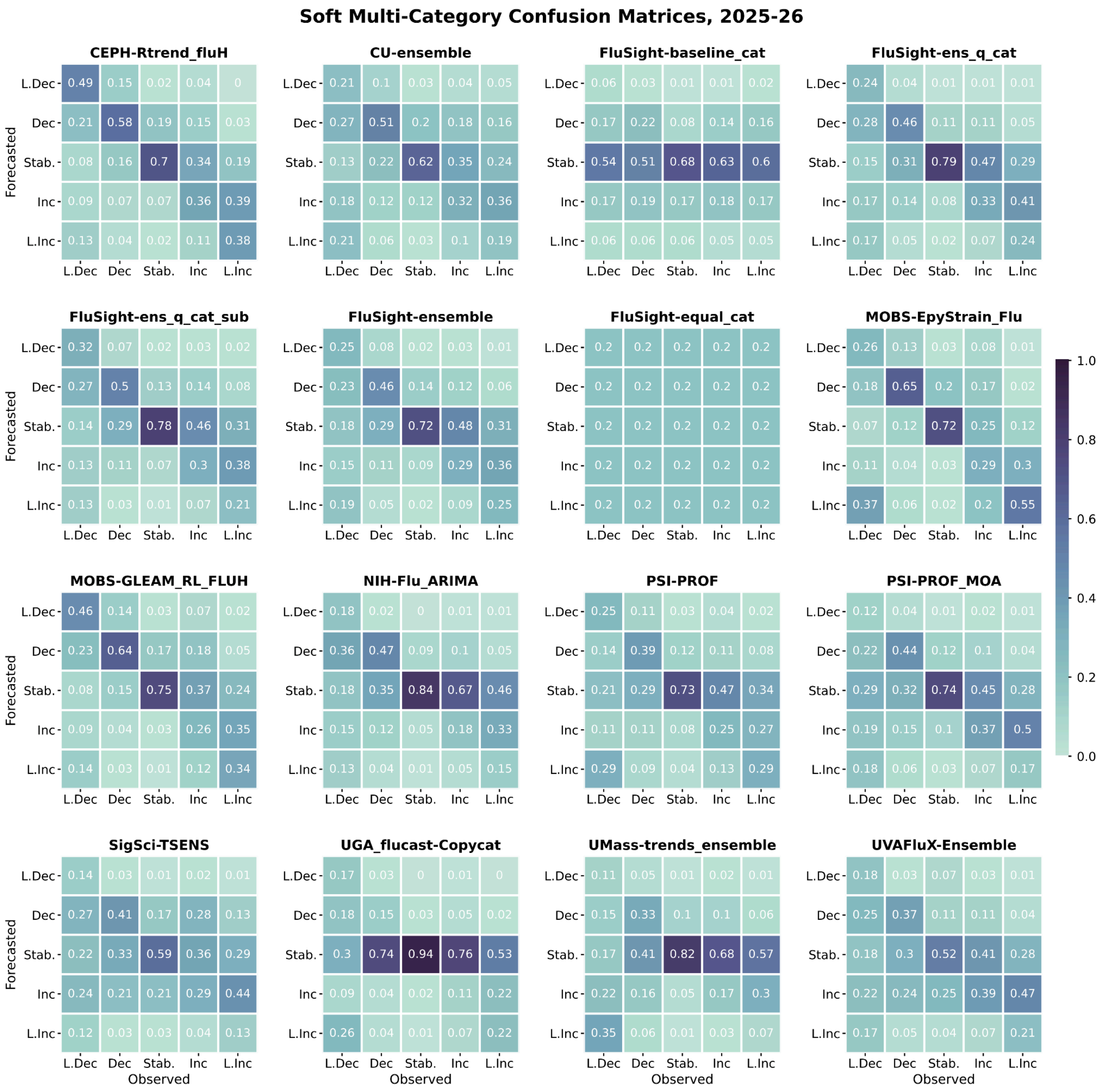

*Figure S8: Confusion Matrices for all teams during the 2025–26 flu season.*

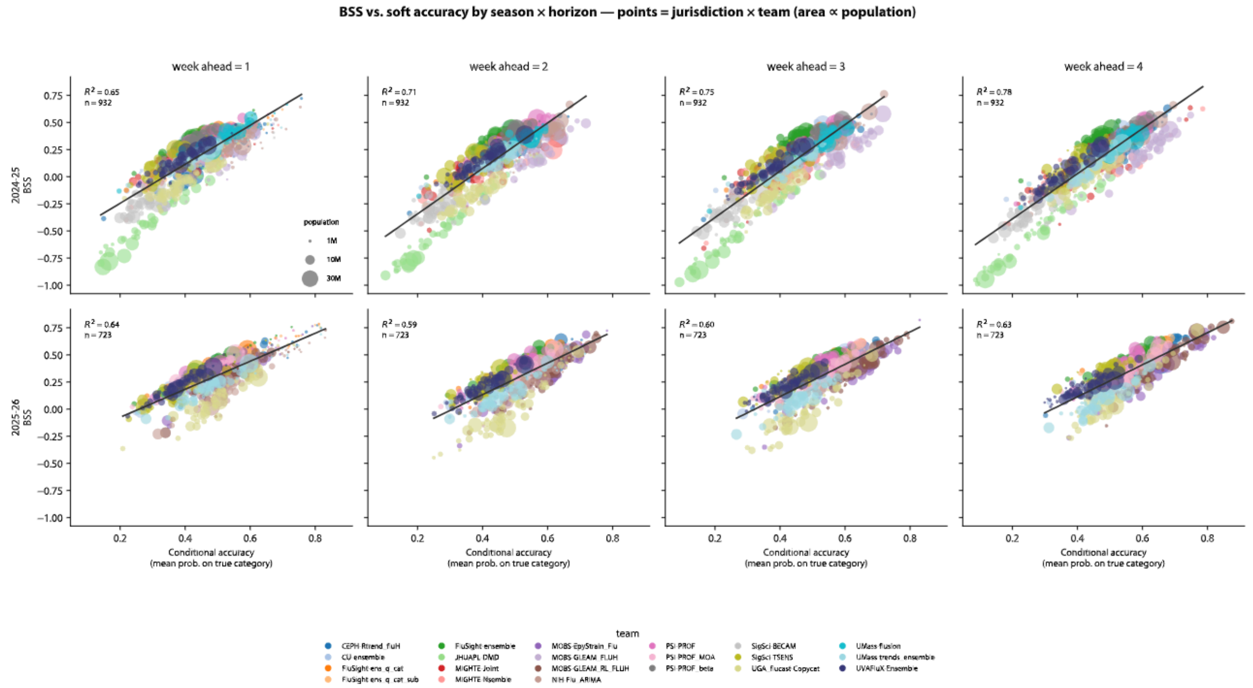

***Figure S9: BSS vs. soft accuracy (mean probability assigned to the true category) by season and horizon.*** *Each point represents a jurisdiction×team pair, with marker area proportional to jurisdiction population and color indicating team. Black lines show the linear fit within each panel, with corresponding* $R^2$ *values reported. The two reference models FluSight-equal_cat and FluSight-baseline_cat were removed from the correlation.*

### *4*. Sensitivity analysis

#### *4.1 Effect of the reference model*

We evaluate the Brier skill scores by the observed category (see section S5) for each model using the equal probability model *FluSight-equal_cat* and the FluSight baseline converted to category model *FluSight-baseline_cat* as reference models. The two references penalize models differently by category. For the stable category, the baseline reference model assigns high probability to the stable category, making it harder to outperform. Therefore, all models show higher skill when evaluated against the equal category reference model than against the baseline reference model (Fig. S10). In contrast, for all other categories, the baseline reference model represents a comparatively weaker benchmark, and the models therefore generally achieve higher skill scores compared to the equal probability model (Fig. S10). However, even with category-specific differences, the aggregated Brier skills scores (as defined in Eq. S5) obtained using the two reference models are strongly correlated and in general the BSS computed relative to the baseline reference model is higher than the BSS computed relative to the equal-probability reference model (Fig. S11). Model rankings are largely preserved across the two references, indicating that the qualitative conclusions in the main text are not sensitive to the choice of reference models studied here.

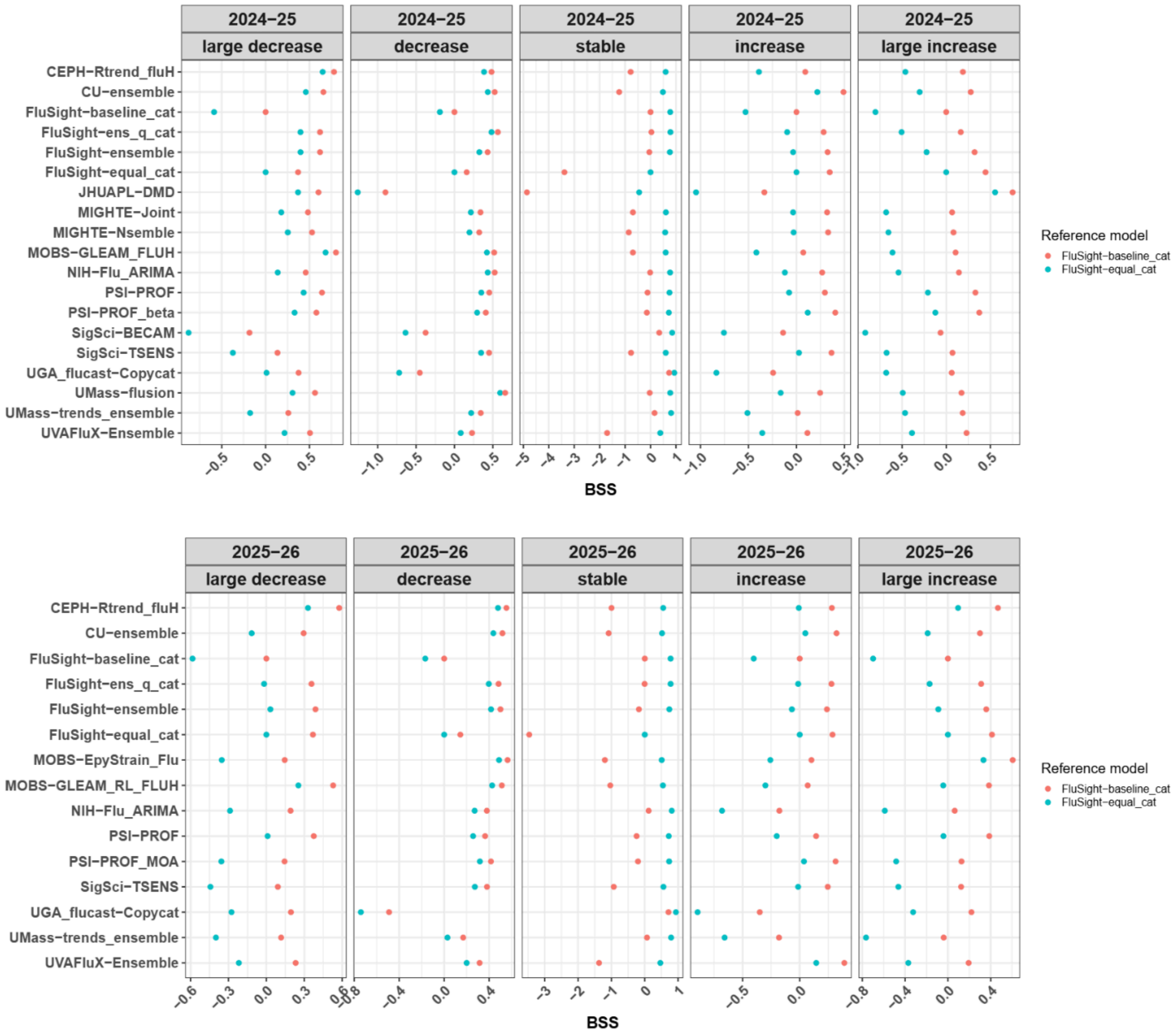

**_Figure S10: Brier Skill Score (BSS) for all models by observed categories for the 2024–25, and 2025–26 seasons._** _The colored points represent the BSS with different reference models._

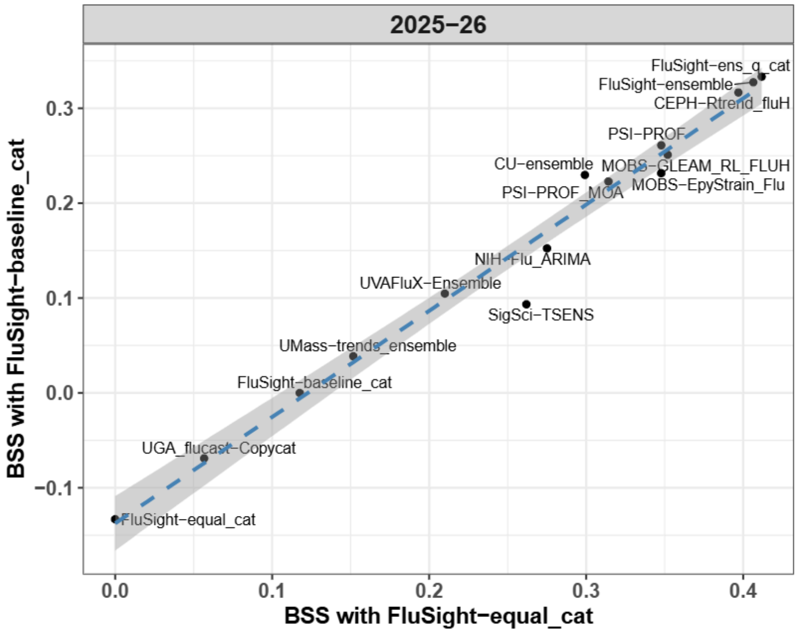

***Figure S11: BSS for each model relative to different reference models.*** *The x-axis shows BSS computed relative to the FluSight-equal_cat reference model, while the y-axis shows BSS computed relative to the FluSight-baseline_cat reference model.*

#### *4.2 Effect of scoring rule: comparing the ranked probability score*

The main text analysis uses the Brier Score (BS) and Brier Skill Score (BSS) as the primary scoring metrics. Here we complement that analysis with the ranked probability score (RPS), which respects the ordering of the trend categories by penalizing probability mass more heavily the further it sits from the observed category (see Methods). Note that the bounds of the BS and RPS as defined in the Methods section of the main text range from 0 (perfectly correct forecast) to 1 (perfectly incorrect forecast).

Tables S7 and S8 illustrate the key difference between the RPS and BS. When the observed category, $o$, is fixed and the forecasted category varies, with all probability mass placed on a single category (Table S7, observation = *large increase*), RPS declines as the forecast moves toward the truth, while the BS treats every incorrect category identically. However, when the forecast is fixed and completely uncertain (equal probability per category) and the observation varies (Table S8), the BS is constant across observed categories but the RPS depends on the observed category. RPS is lowest for *stable* and highest for the two extremes, since under a uniform forecast the average distance from the misplaced mass to the truth is smallest when the truth is in the middle of the ordering.

*Table S7: **RPS and BS for different forecasted categories when the observed category is large increase.** Columns show the observed indicator o, the forecasted probability $P_m$, and their cumulative versions $F_o$ and $F_m$. RPS decreases as the forecasted category approaches the observed category, while BS is constant across all incorrect categories.*

| Forecasted category | $o$ (obs) | $P_m$ (forecast) | $F_o$ (cumul. obs) | $F_m$ (cumul. forecast) | RPS | BS |
|---|---|---|---|---|---|---|
| *large decrease* | [0,0,0,0,1] | [1,0,0,0,0] | [0,0,0,0,1] | [1,1,1,1,1] | 1 | 1 |
| *decrease* | [0,0,0,0,1] | [0,1,0,0,0] | [0,0,0,0,1] | [0,1,1,1,1] | 0.75 | 1 |
| *stable* | [0,0,0,0,1] | [0,0,1,0,0] | [0,0,0,0,1] | [0,0,1,1,1] | 0.5 | 1 |
| *increase* | [0,0,0,0,1] | [0,0,0,1,0] | [0,0,0,0,1] | [0,0,0,1,1] | 0.25 | 1 |
| *large increase* | [0,0,0,0,1] | [0,0,0,0,1] | [0,0,0,0,1] | [0,0,0,0,1] | 0 | 0 |

*Table S8: **RPS and BS for the uniform reference model forecast ($P_r$= [0.2,0.2,0.2,0.2,0.2]) by observed category.** Columns show the observed indicator o, the forecast probability $P_r$, and their cumulative versions $F_o$ and $F_r$. BS is constant at 0.4 across all observed categories, while RPS is lowest for stable and highest for the extreme categories.*

| Observed category | $o$ (obs) | $P_r$ (uniform forecast) | $F_o$ (cumulative obs) | $F_r$ (cumulative forecast) | RPS | BS |
|---|---|---|---|---|---|---|
| *large decrease* | [1,0,0,0,0] | [0.2,0.2,0.2,0.2,0.2] | [1,1,1,1,1] | [0.2,0.4,0.6,0.8,1] | 0.30 | 0.4 |
| *decrease* | [0,1,0,0,0] | [0.2,0.2,0.2,0.2,0.2] | [0,1,1,1,1] | [0.2,0.4,0.6,0.8,1] | 0.15 | 0.4 |
| *stable* | [0,0,1,0,0] | [0.2,0.2,0.2,0.2,0.2] | [0,0,1,1,1] | [0.2,0.4,0.6,0.8,1] | 0.10 | 0.4 |
| *increase* | [0,0,0,1,0] | [0.2,0.2,0.2,0.2,0.2] | [0,0,0,1,1] | [0.2,0.4,0.6,0.8,1] | 0.15 | 0.4 |
| *large increase* | [0,0,0,0,1] | [0.2,0.2,0.2,0.2,0.2] | [0,0,0,0,1] | [0.2,0.4,0.6,0.8,1] | 0.30 | 0.4 |

The results above also have implications for the skill score that uses the RPS and the corresponding reference models. To assess model ($m$) performance against a reference model ($r$) using RPS, we define the Ranked Probability Skill Score for forecast target of a given location ($l$), forecast week ($t$), and number of weeks ahead ($a$), $(l, t, a)$ as

$$RPSS(m, (l, t, a)) = 1 - \frac{RPS(m,(l,t,a))}{RPS(r,(l,t,a))}. \quad \text{(S1)}$$

$RPSS$ ranges between $-4$ and 1. Where an RPSS of 1 represents a perfect forecast, a score of 0 means that it is equivalent to the the reference model (an equal probability across all scores), and -4 is the lower bound assuming the same reference model (see Section S5). Note that changing the reference model changes the bounds. A consequence of the target-dependent baseline RPSS shown in Table S8 is that the baseline is hardest to outperform on *stable* weeks, where its RPS is lowest. Table S9 compares the scores and skills of the BS and RPS across a set of forecast scenarios.

***Table S9: Illustrative examples of skill measures for different forecasting scenarios.***

| Forecast Scenario | Observed category | $P_m$ (forecast) | BS | BSS | RPS | RPSS |
|---|---|---|---|---|---|---|
| Perfect forecast | *stable* | [0,0,1,0,0] | 0 | 1 | 0 | 1 |
| Better than baseline forecast | *stable* | [.1,.1,.4,.2,.2] | 0.23 | 0.425 | 0.0625 | 0.375 |
| Baseline | *stable* | [.2,.2,.2,.2,.2] | 0.4 | 0 | 0.1 | 0 |
| Very poor forecast | *stable* | [0,0,0,0,1] | 1 | -1.5 | 0.5 | -4 |

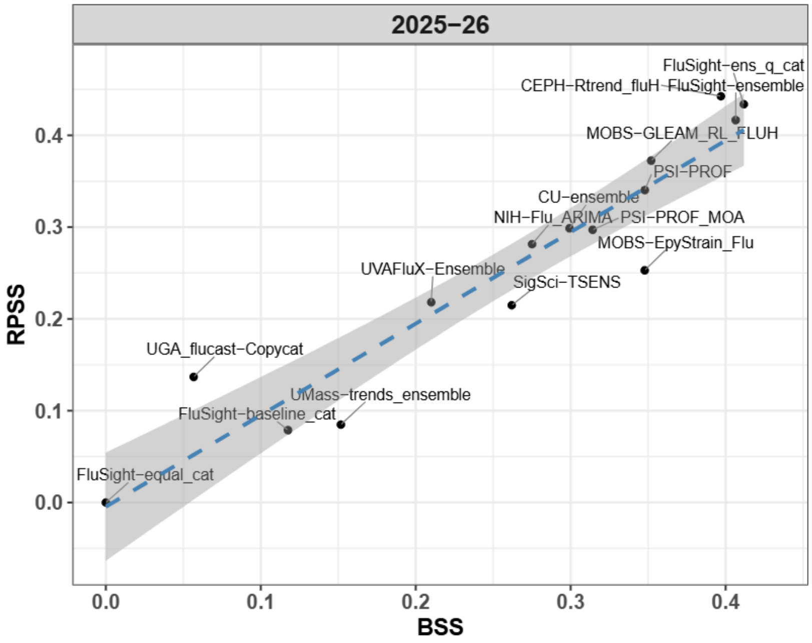

***Figure S12: Aggregated Brier Skill Scores (BSS) and Ranked Probability Skill Scores (RPSS) for all eligible models during 2025–26, as defined in Equation S5 and S6.*** *Compares aggregated BSS and RPSS for all eligible models during the 2025–26 season, computed using Equations S5 and S6. Model rankings under the two metrics are largely consistent, indicating that the qualitative conclusions in the main text are not sensitive to the choice of scoring rule.*

#### 5. Properties of skill scores

While the definitions of forecast skill used are guaranteed to lie in $(-\infty, 1]$ for any type of (non-negative) finite scoring rule and any reference model selection, BSS and RPSS with the *FluSight-equal_cat* reference specifically have a much narrower range of possible values. Below we derive these bounds and the corresponding aggregation properties.

##### *Range of possible values for BSS*

The *FluSight-equal_cat* reference model $r$ outputs the probability distribution $P_r = [1/5, ..., 1/5]$ for every prediction target, assigning equal probability to each of the $K = 5$ categories. For this model, the Brier Score is constant across all observed categories (across all targets $(l, t, a)$) and given by:

$$BS(r) = \frac{1}{2}\sum_{j=1}^{5}\left(\frac{1}{5} - o_j\right)^2 = 0.4,$$

where $o_j$ is the indicator for the observed category. For a forecast target $(l, t, a)$, the Brier Skill Score for a model $m$ is

$$BSS(m, (l, t, a)) = 1 - \frac{BS(m,(l,t,a))}{BS(r,(l,t,a))} = 1 - \frac{BS(m,(l,t,a))}{0.4}. \tag{S2}$$

Recall that $BS(m, (l, t, a))$ is already scaled to be between 0 and 1; thus $BSS(m, (l, t, a))$ must lie between $-1.5$ and 1. The same bounds also apply to aggregate BSS.

##### *Range of possible values for RPSS*

Similarly, the range of possible single-target RPS values for both $m$ and $r$ constrains the possible values of RPSS for $m$. The RPS of $m$ is constructed to lie in $[0, 1]$ as with BS. The RPS of *FluSight-equal_cat* does not have the same value for every target, but will have the same RPS for any two targets where the observed category was the same. Comparing against pessimistic forecasts for each category (illustrated for the *stable* category in Table S9), we find that RPSS lies in $[-4, 1]$ with this baseline. This range also applies to aggregate RPSS values based on their interpretation as a weighted average, below.

##### *Interpreting generic forecast skill aggregations*

Note that aggregate BSS and RPSS are not defined as simple averages of scores for individual predictions. The more complicated aggregations used are analogous to those used in the "relative WIS" metric used in quantile forecast evaluation[3], which was designed in part to (i) ensure that models are not ranked higher simply because they only submitted predictions for an "easier" set of targets, and (ii) avoid over-emphasizing targets where the baseline model has low error/score.

Skill aggregations such as overall forecast skill and forecast skill by category have some generic properties that hold regardless of the underlying score and reference model, and some special properties for BS and for RPS with the *FluSight-equal_cat* reference.

Generic aggregate skill properties:

$$Skill(m,\ S)\ =\ 1\ -\ \frac{\sum_{(l,t,a)\ \in I_{m,r\ \cap S}} Score\ (m,\ (l,t,a))}{\sum_{(l,t,a)\ \in I_{m,r\ \cap S}} Score(r,\ (l,t,a))} \quad \text{(S3)}$$

$$=\ \sum_{(l,t,a)\ \in\ I_{m,r\ \cap S}} \frac{Score\ (r,\ (l,t,a))}{\sum_{(l',t',a')\ \in I_{m,r\ \cap\ S}} Score(r,\ (l',t',a'))} Skill(m,\ (l,t,a)) \quad \text{(S4)}$$

that is, aggregate skill on a subset can be seen as a weighted average of skill values for individual predictions, with weights proportional to the reference model's error/score on those predictions. The same property also holds for overall aggregate skill score.

Similarly, we can relate overall aggregate skill to various breakdowns (e.g., aggregate skill by ahead) using weighted averages, with weights proportional to the reference model score *sum* within each subset:

$$Skill(m)\ =\ \sum_{i=1}^{n} \frac{\sum_{(l,t,a)\in I_{m,r}\cap S_i} Score(r,(l,t,a))}{\sum_{i'=1}^{n}\sum_{(l,t,a)\in I_{m,r}\ \cap\ S_{i'}} Score(r,(l,t,a))} Skill(m, S_i),$$

where $S_1 \ldots S_n$ partition the set of all prediction targets into $n$ subsets, e.g., $S_1$ is the set of all one-ahead targets, $S_2$ is the set of all two-ahead targets, etc.

##### *Special properties of BSS and RPSS aggregations with FluSight-equal_cat reference model*

The limited set of possible BS and RPS values for this reference enables stronger statements to be made about aggregations in these cases. The aggregated Brier Skill Score ($BSS(m)$) is equivalent to the *unweighted* average of individual BSS values across all targets:

$$BSS(m)\ = \frac{1}{|I_{m,r}|} \sum_{(l,t,a)\in I_{m,r}} BSS(m, (l, t, a)). \quad \text{(S5)}$$

However, this property does not hold for the aggregated Ranked Probability Skill Score ( $RPSS(m)$):

$$RPSS(m) \neq \frac{1}{|I_{m,r}|} \sum_{(l,t,a)\in I_{m,r}} RPSS(m, (l, t, a)).$$

This is because the denominators of individual RPS values vary across targets. However, a weaker property holds for (both BSS and) RPSS on this reference model: the skill *conditioned on a given category* is the same as the average of individual skills across targets *of that category*. That is:

$$RPSS(m, S_c) = \frac{1}{|I_{m,r} \cap S_c|} \sum_{(l,t,a) \in I_{m,r} \cap S_c} RPSS(m, (l, t, a)),$$

where $S_c$ is the set of targets with eventually-observed rate-trend category $c$. This is because the reference model score depends on $(l, t, a)$ only through the observed category $c$. That is, for a fixed observed category, the denominator is common across all targets.

This also means that we can interpret overall RPSS as a category-weighted average over individual RPSS scores:

$$RPSS(m) = \frac{\sum_c \sum_{(l,t,a) \in I_{m,r} \cap S_c} w_c RPSS(m, (l,t,a))}{\sum_c \sum_{(l,t,a) \in I_{m,r} \cap S_c} w_c},$$

where the values of $w_c = RPS(r, S_c)$ are 0.30, 0.15, 0.10, 0.15, and 0.30 for $c$ values of "large decrease", "decrease", "stable", "increase", and "large increase", respectively. Note that this weighted average is based on individual RPSS values, which also incorporate the weight $w_c$ internally, rather than raw RPS values. Relating more directly to raw RPS values, we have:

$$RPSS(m) = 1 - \frac{\sum_{(l,t,a) \in I_{m,r}} RPS(m, (l,t,a))}{\sum_c |I_{m,r} \cap S_c| \, w_c} \quad \text{(S6)}$$

##### *Implications for interpreting forecast skill aggregations*

In the case of relative WIS on count-scale quantile forecasts (skill based on $Score$ being weighted interval score) with the *FluSight-baseline* quantile forecaster, we expect baseline WIS to be higher during peaks and in more populous locations, so this method of aggregation can be seen as emphasizing individual relative WIS contributions from these situations. This avoids the problem that simple arithmetic or geometric averages would have: overemphasizing predicting noise during low and flat periods, which are less critical. However, in the case of BS and RPS, the scale of possible scores has already been somewhat standardized across times and locations. Coupled with the choice of the *FluSight-equal_cat*, BSS aggregations are actually equivalent to simple averages, and RPSS aggregations are equivalent to weighted averages with weights determined by the observed category, as seen above.

#### 6. The impact of backfill in the surveillance data

Backfill in the data available at the time of forecast generation (Wednesday) was present across both seasons and varied in both magnitude and frequency. Although reporting was mandatory throughout, delays and corrections were common for the most recent data point. In addition, a shift from daily to weekly reporting in November 2024 altered the timing and structure of these revisions. In the 2024–25 and 2025–26 seasons, preliminary weekly totals were released each Wednesday for the week ending the prior Saturday, with more complete values released on

Friday and in later updates. In all cases, the most recent data point at forecast time was subject to revision.

Fig S13 shows per-capita backfill and  signed-log backfill (defined as the final values reported on 2026-07-01 minus the values reported at the time the forecasts were made) by location for the 2024–25 and 2025–26 seasons. Positive backfill dominates, indicating that final values generally exceed initial reports. However, negative revisions are also observed, with several jurisdictions exhibiting negative median backfill during 2024–25 (e.g., CT, MN, OH, and WA). As summarized in Fig S13 and Table S10, the 2024–25 season exhibits larger average absolute backfill and a higher fraction of negative revisions. In contrast, the 2025–26 season shows a slightly higher overall fraction of jurisdiction–weeks with any revision to the most recent data point.

We also examined associations between the mean *FluSight-ensemble* overall BSS for each location and four backfill metrics: (i) percentage of week–location pairs with negative backfill, (ii) percentage of weeks with any backfill, (iii) mean absolute per-capita backfill, and (iv) mean negative per-capita backfill. Table S11 illustrates our findings using the mean absolute per-capita backfill metric.  Across both seasons, we found no consistent association between forecast performance and mean absolute per-capita backfill, nor the other backfill metrics, neither overall nor for any 1- to 4-week-ahead horizon.

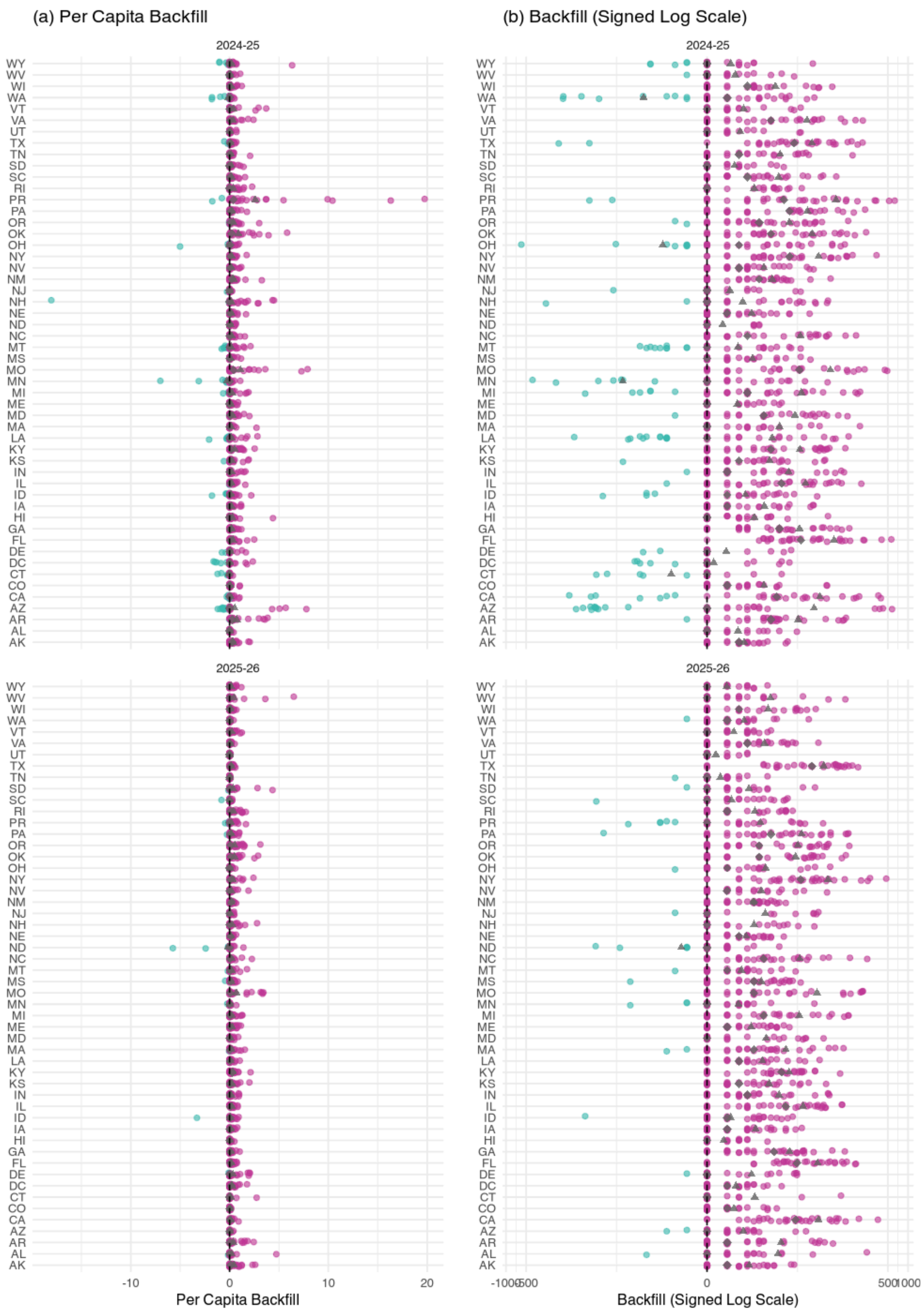

***Figure S13: Per capita and signed-log representation of the backfill pattern for each location during the 2024–25 (top) and 2025–26 (bottom) seasons.*** *Backfill is defined as the difference between the final values as of 2026-07-01 and the values at the time the forecasts were made, with green and purple indicating negative and positive backfill, respectively. Median and mean values are shown using gray diamond and triangle markers. The left and right panels display backfill in raw counts and per-capita values, respectively.*

***Table S10: Season-level summary of backfill.*** *Statistics are computed across all jurisdictions–week pairs for the 2024–25 and 2025–26 seasons.*

| Season | Mean \|Backfill\| | Mean \|Backfill\| (per 100k) | % Negative Backfill | % of Non-zero Backfill |
|---|---|---|---|---|
| 2024–25 | 19.6 | 0.367 | 6.6 | 62.8 |
| 2025–26 | 12.5 | 0.228 | 1.8 | 63.3 |

***Table S11: Pearson correlation between the mean Brier Skill Score of the FluSight-ensemble model for each location and the mean absolute per-capita backfill in that location.*** *Correlations are shown for 1–4 week-ahead forecasts (and overall) during the 2024–25 and 2025–26 seasons. P-values are shown in parentheses. No statistically significant correlations ($p < 0.05$) were found.*

| **Week-ahead** | **2024–25** | **2025–26** |
|---|---|---|
| 1 | -0.07 (0.61) | 0.15 (0.24) |
| 2 | -0.18 (0.21) | -0.06 (0.61) |
| 3 | -0.16 (0.27) | -0.20 (0.12) |
| 4 | 0.01 (0.95) | -0.25 (0.05) |
| Overall | -0.13 (0.35) | -0.07 (0.58) |

#### *6.1 Impact on observed categories*

To assess the impact of surveillance data revisions on categorical trend labels, we tracked how the assigned category for each jurisdiction-week pair changed across successive weekly data releases. Fig S14 shows the percentage of categories matching their final form as a function of weeks since data release, stratified by horizon. Horizon 0 labels were the most affected data revisions in both seasons, starting at approximately 81% agreement with the final label right after data release and taking over 20 weeks to reach 95% in 2024–25. Longer horizons stabilized faster, horizons 2 and 3 reached 95% agreement within roughly 5 weeks in both seasons. This is consistent with the horizon 0 category depending on the most recently reported admission rate, which is the value most subject to revision. In 2025–26, stabilization was generally faster across all horizons, with horizons 1–3 reaching near-complete agreement within approximately 5 weeks. However, horizon 0 still lagged, starting near 77% and not reaching 95% until approximately 10 weeks after data release.

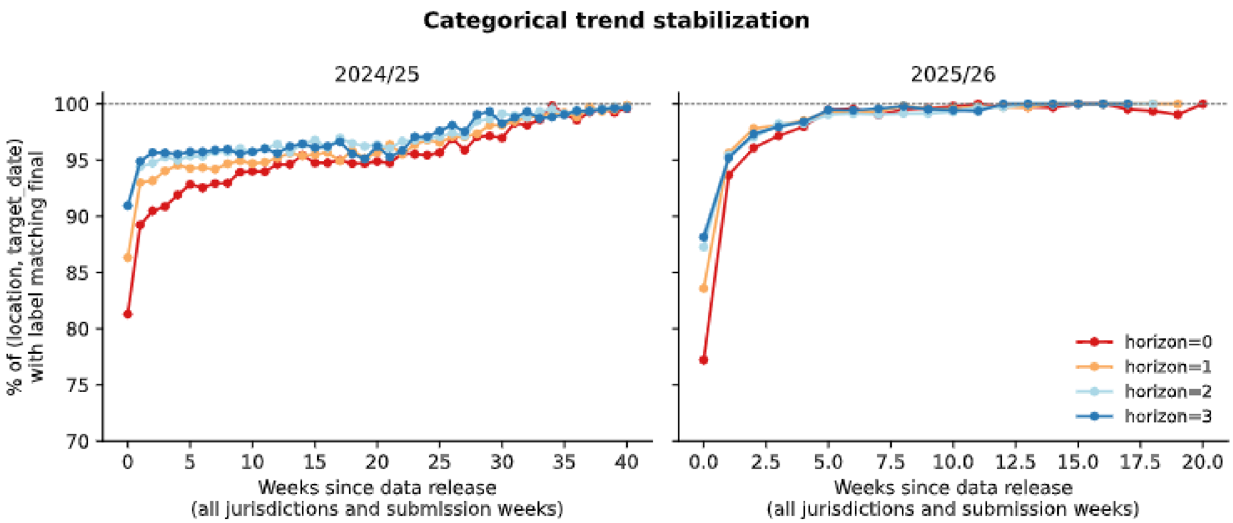

***Figure S14:*** *Each curve shows the percentage of (location, date) pairs whose categorical label — recomputed from an archived surveillance snapshot — agrees with the current label, as a function of the snapshot's age in weeks relative to the target week.*
